# Enhancing Privacy-Preserving Deployable Large Language Models for Perioperative Complication Detection: A Targeted Strategy with LoRA Fine-tuning

**DOI:** 10.1101/2025.06.11.25329235

**Authors:** Shaowei Gao, Xu Zhao, Lihui Chen, Junrong Yu, shuning Tian, Huaqiang Zhou, jingru Chen, Sizhe Long, Qiulan He, Xia Feng

**Author notes:** Corresponding Authors: Xia Feng, MD, PhD Department of Anesthesiology, First Affiliated Hospital of Sun Yat-sen University, Guangzhou, China Qiulan He, MD, PhD Department of Anesthesiology, First Affiliated Hospital of Sun Yat-sen University, Guangzhou, China. Shaowei Gao, MD, PhD Department of Anesthesiology, First Affiliated Hospital of Sun Yat-sen University, Guangzhou, China. These authors contributed equally to this work Affiliations (Shaowei Gao, Xu Zhao, Lihui Chen).

## Abstract

Perioperative complications represent a major global health concern affecting millions of surgical patients annually, yet manual detection methods suffer from significant under-reporting (27%) and misclassification rates. Clinical deployment of large language models (LLMs) for automated complication detection faces substantial barriers including data sovereignty concerns, computational costs, and performance limitations in locally deployable models. Here we show that targeted prompt engineering combined with Low-Rank Adaptation (LoRA) fine-tuning can transform smaller open-source large language models (LLMs) into expert-level diagnostic tools for perioperative complication detection. We conducted a dual-center validation study and developed a comprehensive framework enabling simultaneous identification and severity grading of 22 distinct perioperative complications. Initial evaluations revealed that state-of-the-art models consistently outperformed human experts, particularly reasoning models. AI models maintained consistent performance across varying document complexity, whereas human performance declined with increasing clinical documentation length. Our targeted strategy, which decomposed comprehensive detection into focused single-complication assessments, significantly improved smaller model capabilities. Combined with LoRA fine-tuning, the 4B parameter model’s F1 score increased from 0.18 to 0.55, approaching human expert performance (F1=0.526), while the 8B model achieved F1>0.61. These results demonstrate that optimized smaller models can achieve expert-level diagnostic accuracy while enabling local deployment with preserved data sovereignty, offering a practical solution for resource-limited healthcare institutions to implement automated perioperative complication screening.

## Introduction

Perioperative complications represent a major global health concern, with over 230 million surgical procedures performed worldwide annually and associated mortality rates of 1-4%^1,2^. Beyond mortality risk, complications can lead to reduced functional independence and poorer long-term survival. The World Health Organization has recognized these complications as a global medical issue, linking them to patient suffering, lower quality of life, and significant healthcare costs^3^.

Timely and accurate identification of perioperative complications is vital for effective patient management, quality improvement, and surgical outcome assessment^4,5^. However, current clinical practice relies heavily on manual identification and documentation, methods fraught with significant drawbacks. Studies found 27% of complications, including serious events like occluded vascular grafts and bile leakage, missing from prospective registries^6^. Additionally, approximately 10% of reported events were misclassified, underscoring the prevalence of under-reporting and misclassification. Manual detection is not only error-prone but also time-consuming and labor-intensive, with inconsistencies that compromise data reliability essential for quality improvement and evidence-based practices^5^.

Large Language Models (LLMs), built upon the transformer architecture^7^, have significantly advanced natural language processing, demonstrating remarkable capacity to understand, interpret, and generate human language. In medicine, LLMs show promising applications including clinical documentation assistance, medical query answering, and passing medical licensing exams^8–11^. A key strength is their proficiency in converting unstructured text into structured data, making them highly suitable for extracting clinical information from narrative medical records.

Despite their potential, integrating LLMs into routine clinical practice faces several challenges. General-purpose LLMs may produce “hallucinations”—misleading information often due to lack of specialized medical knowledge^12^. Moreover, many powerful LLMs are proprietary cloud services requiring patient data transmission to external servers, raising significant legal and ethical concerns, especially where personal health data export is restricted by law^13^. Deploying large-scale LLMs also demands significant computational resources, posing major barriers for on-site deployment within healthcare institutions^14^. While open-source LLMs provide locally deployable alternatives, they often perform less effectively than larger commercial models, necessitating optimization strategies for smaller models to accurately identify diverse postoperative complications.

The imperative for practical Artificial Intelligence (AI) deployment in perioperative care is driven by global healthcare institutions’ urgent need to improve patient safety given resource constraints and regulatory pressures. A deployable AI solution offering automated screening and 24/7 “second opinions” could revolutionize perioperative quality management, particularly during challenging periods such as night shifts and holiday coverage when staffing may be reduced and expert availability limited.

This study aims to fill this research gap by exploring ways to improve deployable open-source LLMs’ detection of perioperative complications from clinical records. Specifically, we investigate how combining targeted prompt engineering with LoRa fine-tuning can boost performance of open-source models of various sizes (4B, 8B, 14B, and 32B parameters)^15^. Our main hypothesis is that this specialized approach can enhance smaller, local LLMs’ abilities, making them comparable to, or even better than, human experts and larger, proprietary LLMs for this crucial clinical task. This research could significantly improve complication detection accuracy and efficiency while navigating practical challenges of deploying AI in clinical settings.

## Results

### 1. Data and design

To evaluate LLM effectiveness in detecting postoperative complications from clinical records, this study utilized a dual-center design. Data were retrospectively collected from two independent medical centers: Center 1 (n=146 cases) served as the primary dataset for model training and validation, while Center 2 (n=52 cases) provided external validation.

The study cohort included patients undergoing diverse surgical procedures across multiple specialties (Table 1). Center 1 (n=146) had a mean patient age of 57.7±14.3 years and male predominance (65.8%), with surgical cases mainly from gastrointestinal (44.5%), hepatobiliary and pancreatic (32.9%), and urological (11.0%) surgery. Center 2 (n=52) had a mean patient age of 58.2±13.7 years with balanced gender distribution (48.1% male), featuring more diverse surgery including orthopedic (23.1%), head and neck/other (15.4%), and gastrointestinal (15.4%). The median length of hospital stay was 12 days (interquartile range (IQR): 9-15) for Center 1 and 9 days (IQR: 7-14) for Center 2. The most frequently observed complications were paralytic ileus (Center 1: 24.7%; Center 2: 11.5%), organ/space surgical site infections (14.4% vs. 7.7%), infections of unknown source (15.1% vs. 5.8%), and myocardial injury after non-cardiac surgery (13.7% vs. 3.8%). These inter-center differences in demographics, surgical specialties, and complication patterns provided a robust platform for assessing model generalizability.

**Table 1.** Patient Demographics and Clinical Characteristics by Center.

| Table 1. Patient Demographics and Clinical Characteristics by Center |  |  |
| --- | --- | --- |
| Characteristic | Center 1 | Center 2 |
| Number of patients | 146 | 52 |
| Gender, n (%) |  |  |
| Male | 96 (65.8%) | 25 (48.1%) |
| Female | 50 (34.2%) | 27 (51.9%) |
| Age, years (Mean ± SD) | 57.7 ± 14.3 | 58.2 ± 13.7 |
| Surgical specialty, n (%) |  |  |
| Head and neck/other | 4 (2.7%) | 8 (15.4%) |
| Gynecological | 5 (3.4%) | 3 (5.8%) |
| Cardiovascular | 5 (3.4%) | 1 (1.9%) |
| Urological | 16 (11.0%) | 3 (5.8%) |
| Neurosurgery | 3 (2.1%) | 3 (5.8%) |
| Hepatobiliary and pancreatic | 48 (32.9%) | 10 (19.2%) |
| Gastrointestinal | 65 (44.5%) | 8 (15.4%) |
| Thoracic | 0 (0.0%) | 4 (7.7%) |
| Orthopedic | 0 (0.0%) | 12 (23.1%) |
| **Median Length of hospital stay, days (IQR)** | 12 (9-15) | 9 (7-14) |
| **Complications, n (%)** |  |  |
| **Paralytic ileus** | 36 (24.7%) | 6 (11.5%) |
| **Organ/space surgical site infection** | 21 (14.4%) | 4 (7.7%) |
| **Infection of unknown source** | 22 (15.1%) | 3 (5.8%) |
| **Myocardial injury after non-cardiac surgery** | 20 (13.7%) | 2 (3.8%) |
| **Anastomotic leakage** | 15 (10.3%) | 3 (5.8%) |
| **Pneumonia** | 10 (6.8%) | 3 (5.8%) |
| **Postoperative bleeding** | 10 (6.8%) | 2 (3.8%) |
| **Acute kidney injury** | 7 (4.8%) | 5 (9.6%) |
| **Superficial surgical site infection** | 4 (2.7%) | 6 (11.5%) |
| **Arrhythmia** | 7 (4.8%) | 2 (3.8%) |
| **Cardiogenic pulmonary edema** | 7 (4.8%) | 0 (0.0%) |
| **Deep vein thrombosis** | 5 (3.4%) | 2 (3.8%) |
| **Deep surgical site infection** | 3 (2.1%) | 3 (5.8%) |
| **Gastrointestinal bleeding** | 4 (2.7%) | 2 (3.8%) |
| **Delirium** | 5 (3.4%) | 0 (0.0%) |
| **Acute respiratory distress syndrome** | 4 (2.7%) | 1 (1.9%) |
| **Pulmonary embolism** | 3 (2.1%) | 0 (0.0%) |
| **Myocardial infarction** | 2 (1.4%) | 0 (0.0%) |
| **Bloodstream infection** | 1 (0.7%) | 1 (1.9%) |

For each case, we systematically extracted comprehensive clinical information including patient demographics, surgical procedure details, and postoperative findings—medical records, abnormal laboratory, and imaging results. Our methodological framework (Fig. 1) involved systematic prompt engineering and model selection, followed by iterative refinement driven by performance analysis, integrating both comprehensive and targeted strategies, culminating in LoRa fine-tuning of open-source models.

**Figure 1.**
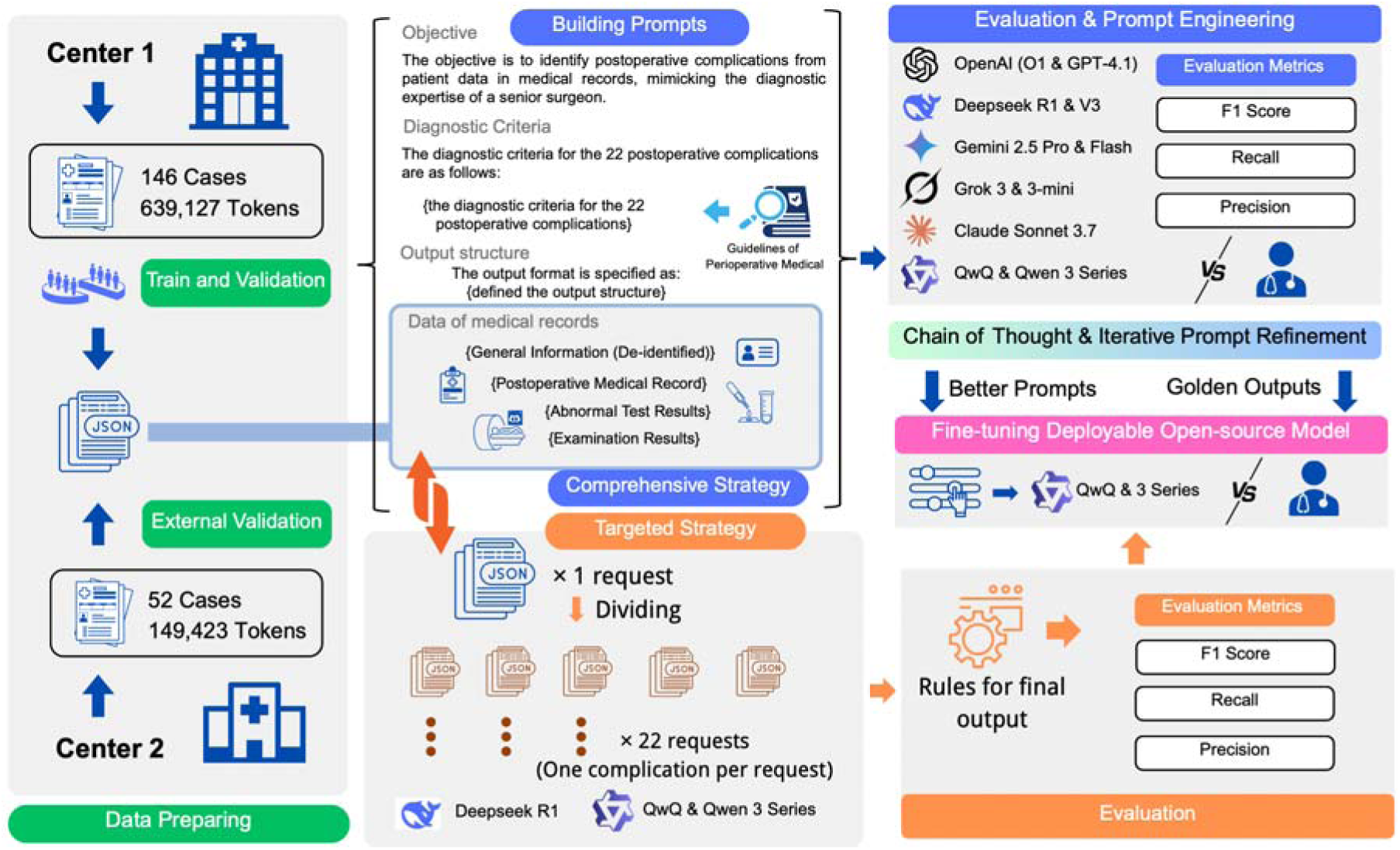
Methodological framework for AI-enhanced perioperative complication detection. The study utilized a dual-center design with data collection from Center 1 (n=146 cases, 639,127 tokens) and Center 2 (n=52 cases, 149,423 tokens). The workflow included: (1) prompt engineering using European Perioperative Clinical Outcome (EPCO) diagnostic criteria, (2) iterative evaluation and optimization across multiple AI models, and (3) comparison of comprehensive versus targeted strategies followed by LoRa fine-tuning of open-source models. Performance assessed using F1 score, recall, and precision metrics with comparative analysis between models and against human expert benchmarks.

### 2. Prompt Development and Initial Evaluations

Following our prompt construction methodology (see Methodology section; Fig. 1, Fig. 2a), we developed a comprehensive prompt framework enabling simultaneous identification and severity grading of 22 distinct perioperative complications—a notable improvement over traditional binary classification. Analysis of prompt token counts (Fig. 2b) revealed considerable heterogeneity between medical centers: Center 1 featured substantially longer clinical documentation (average 6,841.6 tokens) compared to Center 2 (average 5,484.6 tokens), offering a rigorous test for model generalizability.

**Figure 2.**
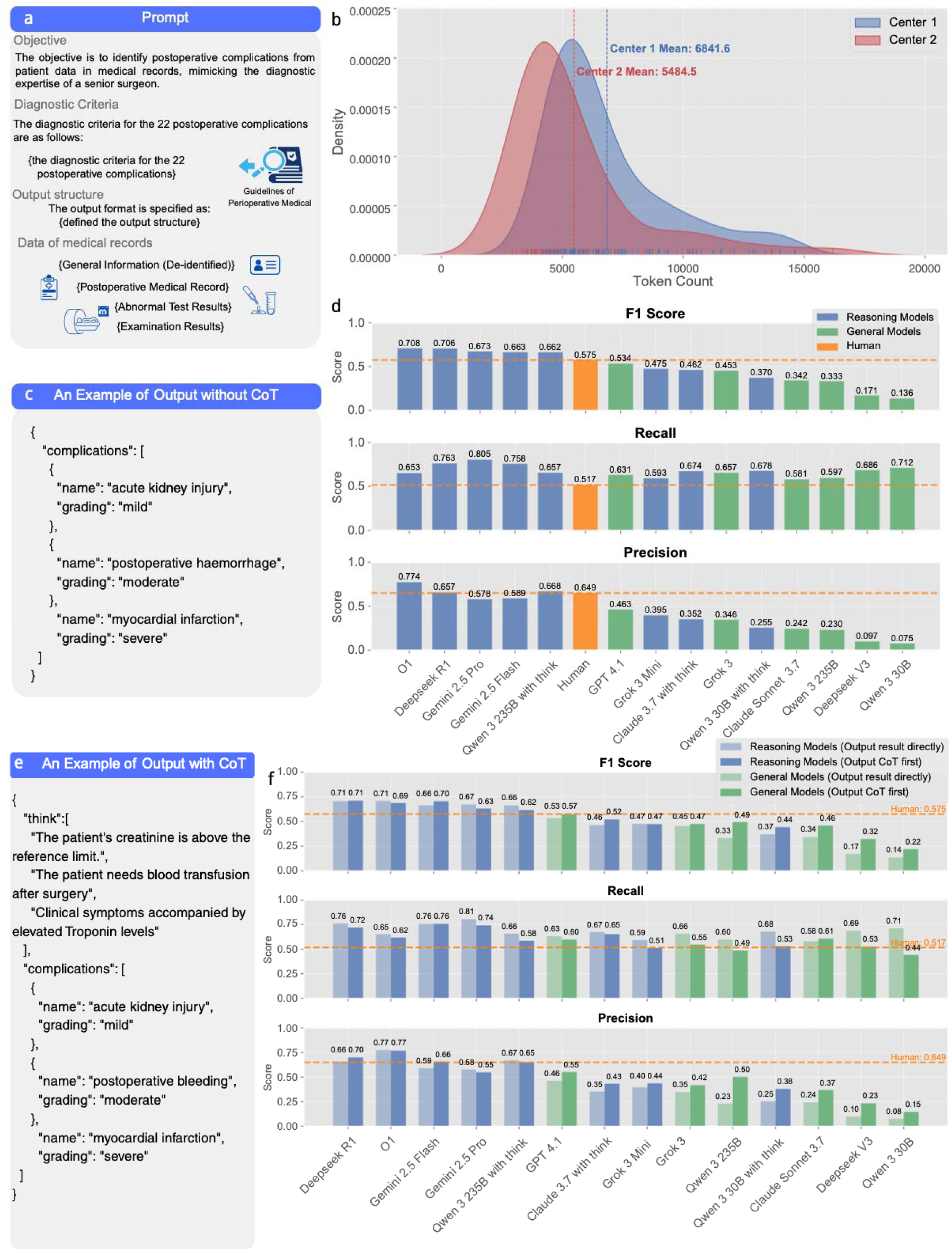
Prompt development and initial model evaluations. (**a**) Comprehensive prompt framework structure including objective definition, European Perioperative Clinical Outcome (EPCO) diagnostic criteria for 22 postoperative complications, structured JavaScript Object Notation (JSON) output format, and clinical data components (general information, postoperative medical records, abnormal test results, and examination results). (**b**) Token count distribution analysis revealing substantial heterogeneity between medical centers, with Center 1 averaging 6,841.6 tokens compared to Center 2’s 5,484.6 tokens per case. (**c**) Example of basic structured JSON output format without Chain-of-Thought (CoT) prompting, showing direct complication identification and severity grading. (**d**) Initial performance evaluation across multiple state-of-the-art language models using micro-averaged metrics (macro-averaged results in Appendix Fig. 1), demonstrating superior performance of reasoning models over general models, with several AI models exceeding human expert benchmarks. (**e**) Example of CoT-enhanced JSON output incorporating diagnostic reasoning through the “think” field, enabling transparent clinical decision-making processes. (**f**) Performance comparison following CoT implementation using micro-averaged metrics (macro-averaged results in Appendix Fig. 2), showing significant improvements in general models while reasoning models maintained consistently high performance across F1 score, recall, and precision metrics.

Initial evaluations of multiple state-of-the-art LLMs using basic structured JavaScript Object Notation (JSON) output format (Fig. 2c) highlighted key performance trends (Fig. 2d, showing micro-averaged metrics; macro-averaged results in Appendix Fig. 1).

Reasoning models consistently outperformed general models, with all models exceeding human performance belonging to the reasoning category. A notable difference in error patterns was observed: AI models generally demonstrated higher recall than precision (more sensitive but less specific), whereas human clinicians exhibited the converse (higher precision, lower recall). AI models were more comprehensive in detection, while human experts adopted more conservative diagnostic approaches.

### 3. Primary prompt engineering

Based on initial evaluation results, we implemented Chain-of-Thought (CoT) prompting to improve diagnostic accuracy and model interpretability^16^. This evolved our structured JSON output from basic diagnostic formats (Fig. 2c) to CoT-enhanced responses (Fig. 2e), integrating reasoning transparency by adding a “think” field where models articulated diagnostic rationale before rendering final conclusions.

CoT implementation revealed distinct performance patterns across model architectures (Fig. 2f showing micro-averaged metrics; macro-averaged results in Appendix Fig. 2). General models demonstrated significant performance enhancements with notable increases in F1 scores, precision, and recall. Conversely, reasoning models showed only marginal gains from explicit CoT prompting. This is consistent with their inherent design: reasoning models possess internal analytical processes, whereas general models derive substantial benefit from externally structured reasoning frameworks. Given consistent performance improvements in general models and maintained high performance in reasoning models, we adopted CoT prompting as the standard approach for all subsequent evaluations.

### 4. Extended analysis and further prompt optimization

To deepen understanding of model behavior and further optimize performance, we undertook extended analysis encompassing stratified evaluation by document complexity and systematic identification of error patterns.

For stratified evaluation, cases were systematically divided into quartiles based on prompt token counts: Q1 (shortest documents, mean: 4,626 tokens), Q2 (short-to-medium, mean: 5,440 tokens), Q3 (medium-to-long, mean: 6,512 tokens), and Q4 (longest and most complex documents, mean: 10,253 tokens). This analysis (Fig. 3a) revealed significant patterns in both human and AI performance. The top-performing model, DeepSeek R1, demonstrated remarkable consistency across document lengths, maintaining consistently high F1 scores irrespective of clinical documentation complexity. In contrast, human performance declined with increasing document length and complexity, suggesting that processing highly complex clinical records is challenging for human experts, potentially owing to cognitive load or fatigue^17,18^, whereas AI models exhibit superior scalability.

**Figure 3.**
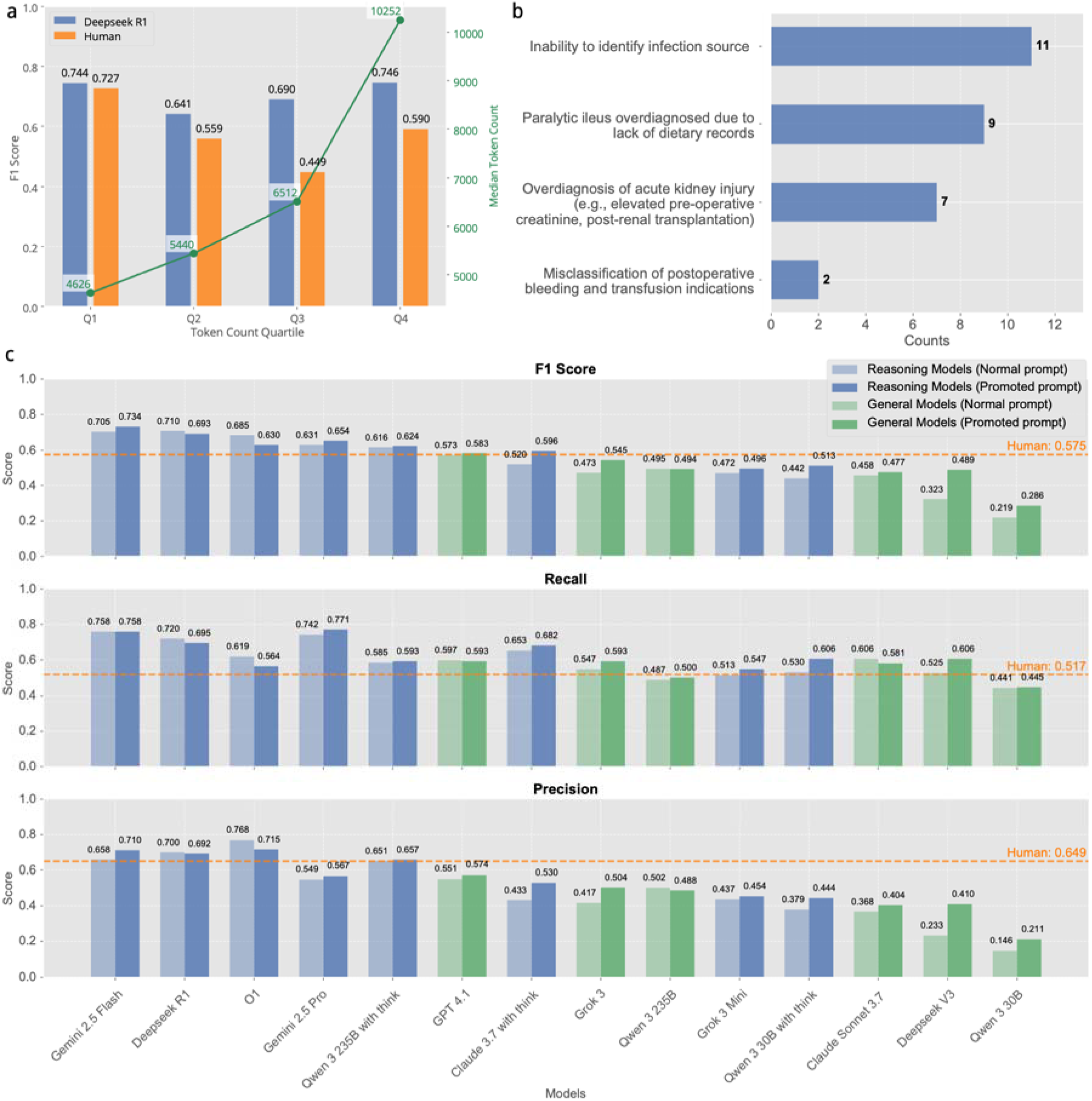
Extended analysis and prompt optimization results. (**a**) Stratified performance evaluation by document complexity, showing F1 scores across quartiles based on token counts (Q1: 4,626 tokens, Q2: 5,440 tokens, Q3: 6,512 tokens, Q4: 10,252 tokens). DeepSeek R1 demonstrates consistent performance across document lengths, while human expert performance declines with increasing complexity. (**b**) Error pattern analysis identifying primary categories of diagnostic failures, including inability to identify infection sources (11 cases), paralytic ileus overdiagnosis due to lack of dietary records (9 cases), overdiagnosis of acute kidney injury (7 cases), and misclassification of postoperative bleeding and transfusion indications (2 cases). (**c**) Performance comparison before and after prompt optimization across multiple models using micro-averaged metrics (macro-averaged results in Appendix Fig. 3), showing substantial improvements in initially lower-performing models. Reasoning models (blue) and general models (green) are compared using normal prompts (lighter colors) versus optimized prompts (darker colors). Orange dashed lines indicate human expert benchmarks for F1 score (0.575), recall (0.517), and precision (0.649).

Systematic review of diagnostic errors identified primary categories of model failures (Fig. 3b), highlighting areas where clinical context interpretation needed refinement. We implemented prompt modifications (see Appendix) incorporating four key enhancements:

**1. Enhanced Diagnostic Specificity**: We incorporated additional clinical exclusion criteria for several complications, such as specifying that acute kidney injury should not be diagnosed following kidney transplantation procedures, and providing explicit baseline creatinine calculation methods when preoperative values were unavailable.
**2. Documentation Clarification**: For paralytic ileus diagnoses, we added guidance regarding incomplete clinical documentation, stipulating that gastrointestinal symptoms (e.g., abdominal distension or vomiting) must be present when flatus or dietary records were incomplete.
**3. Differential Diagnostic Refinement**: We introduced important diagnostic distinctions for postoperative bleeding, specifically excluding cases where postoperative transfusions were administered despite minimal drainage output or pale drainage fluid, directing models to consider these as intraoperative blood loss or pre-existing anemia rather than true postoperative hemorrhage.
**4. Anatomical Precision Enhancement**: For anastomotic leakage, we expanded the definition to explicitly include external leakage, drainage site discharge, and imaging-identified extravasation, providing more comprehensive guidance for this critical complication.

These refinements yielded measurable improvements across most model categories, with more substantial gains observed in models that initially exhibited lower performance (e.g., Qwen 3 30B, Deepseek V3, and Grok 3) (Fig. 3c showing micro-averaged metrics; macro-averaged results in Appendix Fig. 3).

Although our analyses showed that leading commercial models could surpass human clinician performance, their practical implementation in healthcare settings requires addressing crucial concerns about data sovereignty, patient privacy, and computational cost-effectiveness. This necessity guided our investigation into the capabilities of open-source models for perioperative complication detection.

### 5. Open-source AI performance

We systematically evaluated open-source models from diverse manufacturers—including Qwen 3, DeepSeek, QwQ, Gemma, and Mistral—spanning parameter scales from 4B to 671B (Fig. 4a showing micro-averaged metrics; macro-averaged results in Appendix Fig. 4). The quadrant chart reveals a clear correlation between model size and performance: larger parameter counts generally correspond to improved F1 scores. This relationship was not strictly linear, underscoring the significance of architectural design and training methodologies in addition to parameter scaling. Notably, QwQ 32B emerged as a top-performing open-source model, achieving an F1 score of 0.602 in the upper-left quadrant, demonstrating favorable balance between performance and efficiency while surpassing the human clinician benchmark (F1 = 0.575).

**Figure 4.**
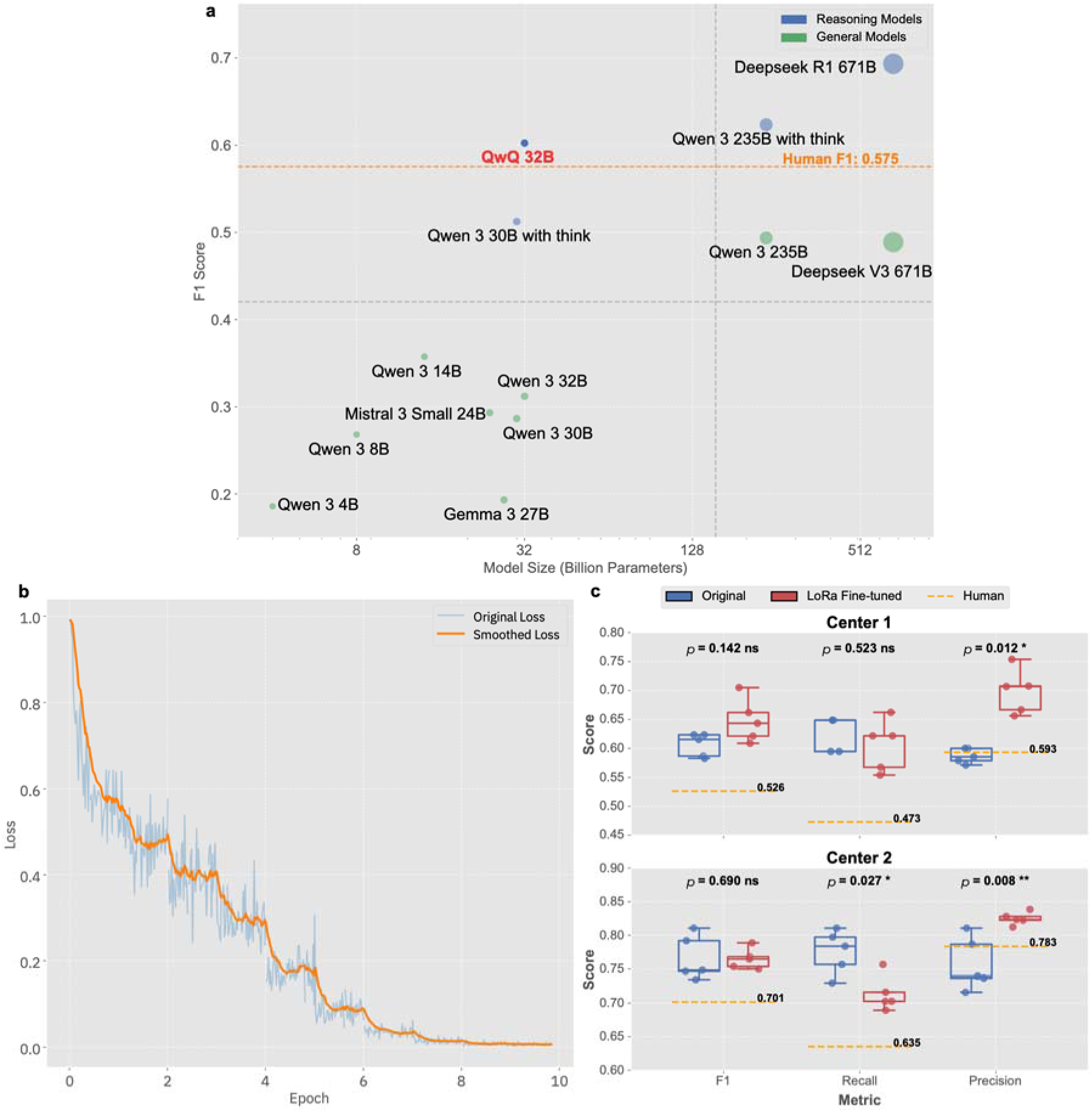
Open-source model performance evaluation and Low-Rank Adaptation (LoRa) fine-tuning results. (**a**) Systematic evaluation of open-source models spanning 4B to 671B parameters using micro-averaged metrics (macro-averaged results in Appendix Fig. 4). Point sizes are proportional to model parameter counts. The quadrant chart demonstrates correlation between model size and performance, with QwQ 32B achieving optimal balance between performance (F1 = 0.602) and efficiency while surpassing human expert benchmark (F1 = 0.575, orange dashed line). Reasoning models (blue) and general models (green) are distinguished by color coding. (**b**) LoRa fine-tuning convergence curves for QwQ 32B, showing original loss (blue) and smoothed loss (orange) trajectories across training epochs, demonstrating stable convergence within reasonable computational budgets. (**c**) Performance comparison before (blue) and after (red) LoRa fine-tuning across F1 score, recall, and precision metrics for both validation datasets. Box plots show distribution of results with statistical significance testing (p-values indicated). Orange dashed lines represent human expert benchmarks for each metric in respective centers.

To further augment open-source model capabilities, we implemented LoRa fine-tuning on QwQ 32B, specifically targeting improved complication detection accuracy. The fine-tuning process (Fig. 4b) demonstrated convergence within reasonable computational budgets. However, performance improvements were modest across multiple evaluation metrics and on both validation data sources, yielding incremental gains rather than dramatic enhancements (Fig. 4c showing micro-averaged metrics; macro-averaged results in Appendix Fig. 5). This limited improvement can be attributed to the model’s already high baseline performance, suggesting it had previously captured much of the clinical knowledge necessary for complication detection.

**Figure 5.**
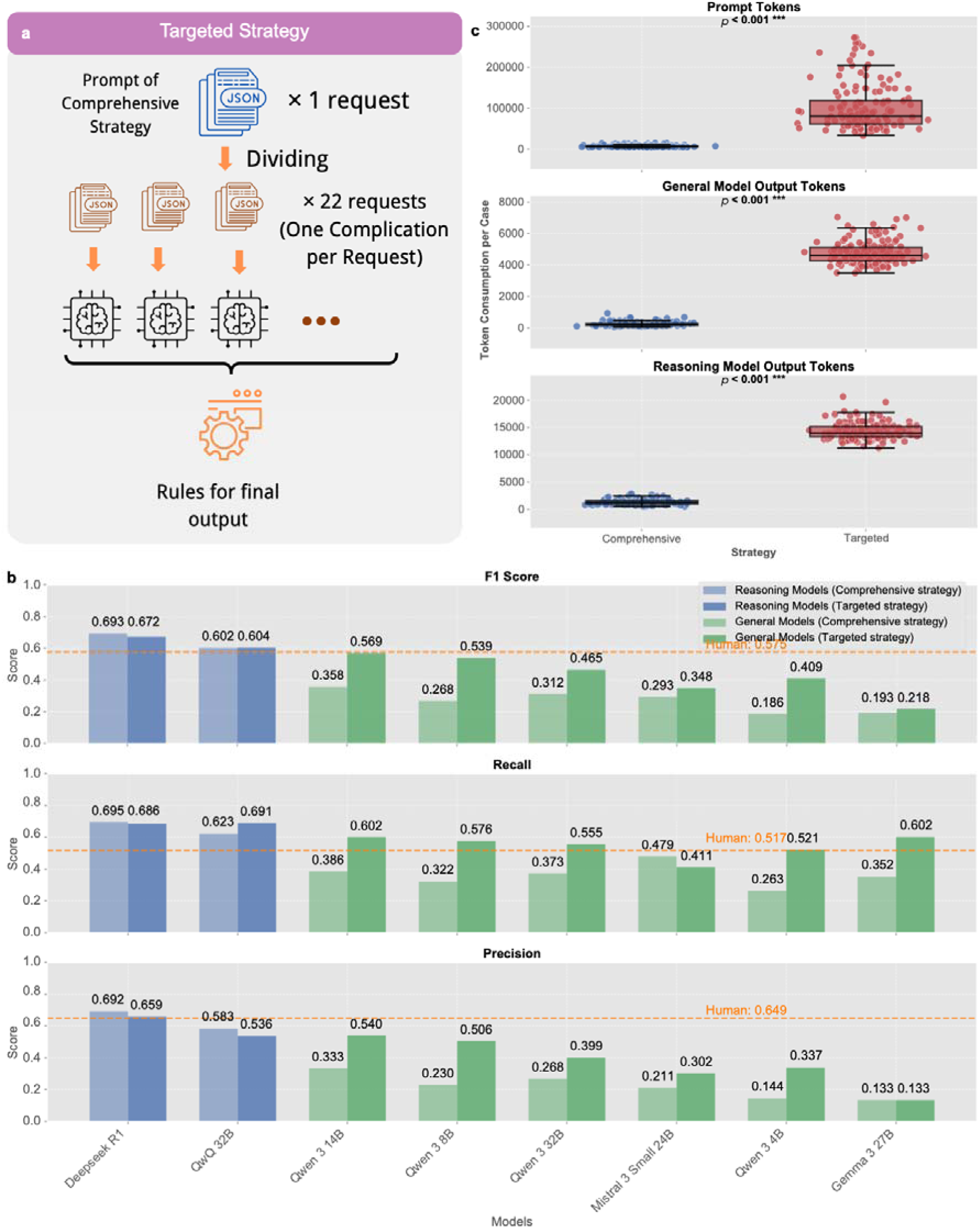
Transition from comprehensive to targeted strategy. (**a**) Schematic illustration of the targeted strategy approach, decomposing the comprehensive detection task (single request evaluating all 22 complications simultaneously) into 22 independent evaluation processes (one complication per request), followed by rules for generating final diagnostic output to ensure clinically coherent and mutually exclusive conclusions. (**b**) Performance comparison between comprehensive strategy (lighter colors) and targeted strategy (darker colors) across F1 score, recall, and precision metrics using micro-averaged results. Reasoning models (light blue and blue) and general models (light purple and purple) demonstrate differential responses to strategy transition, with smaller models showing substantial improvements while larger reasoning models maintain consistent performance. Orange dashed lines indicate human expert benchmarks for each metric. (**c**) Token consumption analysis comparing comprehensive versus targeted strategies across prompt tokens, general model output tokens, and reasoning model output tokens. Box plots show significant increases in computational requirements for the targeted approach (p < 0.001), representing the trade-off between enhanced accessibility for smaller models and increased computational volume.

Despite achieving near-human performance, practical deployment of 32B models exposed critical resource constraints. Clinical documents exceeding 15,000 tokens triggered out-of-memory errors even on high-end 80GB Graphics Processing Unit (GPU) systems, underscoring the urgent need for more efficient alternatives. Smaller models, conversely, grappled with the cognitive complexity of concurrently evaluating 22 distinct complications from extensive clinical narratives. These challenges motivated development of our targeted strategy: decomposing the comprehensive detection task into focused, single-complication assessments.

### 6. Transition from comprehensive to targeted strategy

Our targeted strategy (Fig. 5a) systematically divided the comprehensive detection task into 22 independent evaluation processes, requiring models to evaluate one specific complication per inference call rather than all 22 simultaneously. Each targeted prompt used identical clinical data inputs but was specifically designed to assess one complication type with focused diagnostic criteria and streamlined output requirements. Final diagnostic conclusions adhered to two key rules: (1) if all evaluations yielded negative results, the output was “no postoperative complications”; (2) identification of specific infectious complications automatically excluded a diagnosis of “infection of unknown source” to prevent redundancy. These rules (see “rule to get final output” in Fig. 5a) ensured clinically coherent and mutually exclusive diagnostic conclusions. This decomposition strategy maintained full diagnostic coverage while substantially reducing the cognitive load per inference call. Comparing the two strategies revealed interesting performance patterns (Fig. 5b showing micro-averaged metrics; macro-averaged results in Appendix Fig. 6). Smaller general models showed substantial improvements across all evaluation metrics with the targeted approach. In stark contrast, larger reasoning models exhibited minimal performance changes when shifting strategies. Advanced models such as DeepSeek R1 and QwQ 32B maintained nearly identical performance levels irrespective of strategic approach. This differential response suggests that larger, sophisticated models have sufficient computational capacity for effective comprehensive multi-complication analysis, whereas smaller models benefit considerably from cognitive load reduction inherent in targeted evaluation.

**Figure 6.**
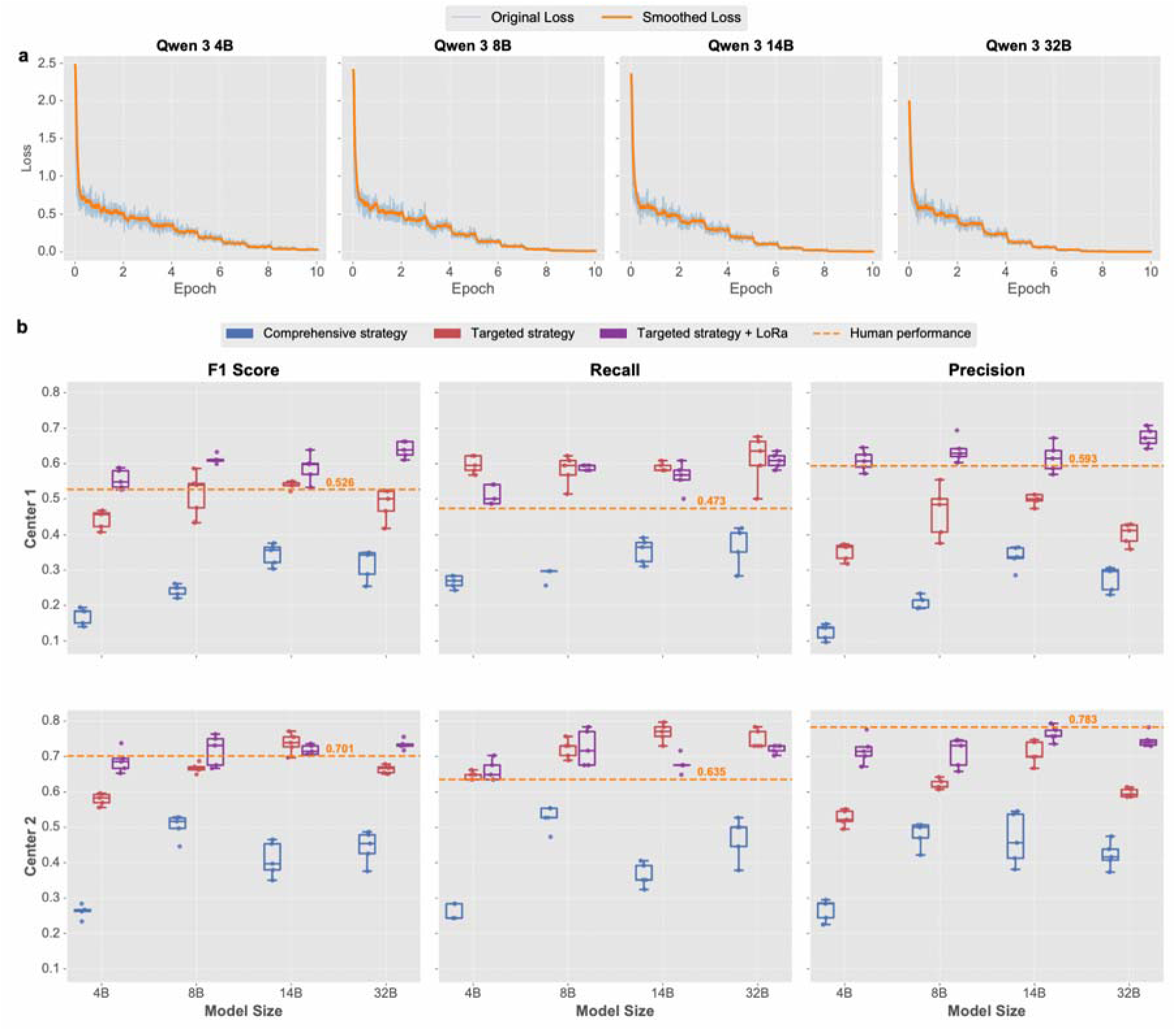
LoRa fine-tuning optimization results for small-scale open-source models. (**a**) Training convergence curves for Qwen 3 models across different parameter scales (4B, 8B, 14B, 32B), showing original loss (blue) and smoothed loss (orange) trajectories during LoRa fine-tuning process. All models demonstrate stable and consistent convergence patterns. (**b**) Comprehensive performance comparison across F1 score, recall, and precision metrics for both Center 1 and Center 2 validation datasets. Box plots illustrate performance distributions for comprehensive strategy (blue), targeted strategy (red), and targeted strategy with LoRa fine-tuning (purple), compared against human expert benchmarks (orange dashed lines). Results demonstrate that LoRa fine-tuning significantly enhances smaller models’ performance, with 4B and 8B parameter models showing substantial improvements and approaching or exceeding human expert performance levels.

However, the targeted strategy substantially increases prompt token consumption per case, requiring multiple focused inferences for comprehensive diagnostic coverage (Fig. 5c). For commercial cloud-based models with per-token charges, this leads to significant cost implications. Conversely, this apparent computational drawback becomes a strategic advantage for local model deployment. In on-premises scenarios with fixed computational costs rather than usage-based charges, the targeted strategy allows smaller models to achieve performance levels previously attainable only by larger, more resource-intensive alternatives. This approach effectively trades increased computational volume for enhanced accessibility, enabling healthcare institutions to deploy smaller models on existing hardware while maintaining diagnostic accuracy.

### 7. Fine tuning of small models

Building on the targeted strategy’s success in optimizing smaller models, we implemented LoRa fine-tuning to further enhance diagnostic capabilities of open-source models across various parameter scales. Our approach focused on four Qwen 3 models, ranging from 4B to 32B parameters, representing a spectrum suitable for diverse clinical environments.

The LoRa fine-tuning process (Fig. 6a) demonstrated consistent convergence patterns across all model sizes with smooth optimization trajectories. Fine-tuning results (Fig. 6b showing micro-averaged metrics; macro-averaged results in Appendix Fig. 7) showed that LoRa adaptation significantly improved model performance across multiple evaluation metrics and on both validation data sources. Statistical significance testing confirmed these improvements across different model sizes and strategies (detailed statistical metrics showed in Table 2 and Appendix Table 1). Smaller models were more responsive to fine-tuning; specifically, the 4B and 8B parameter models exhibited substantial performance improvements.

**Figure 7.**
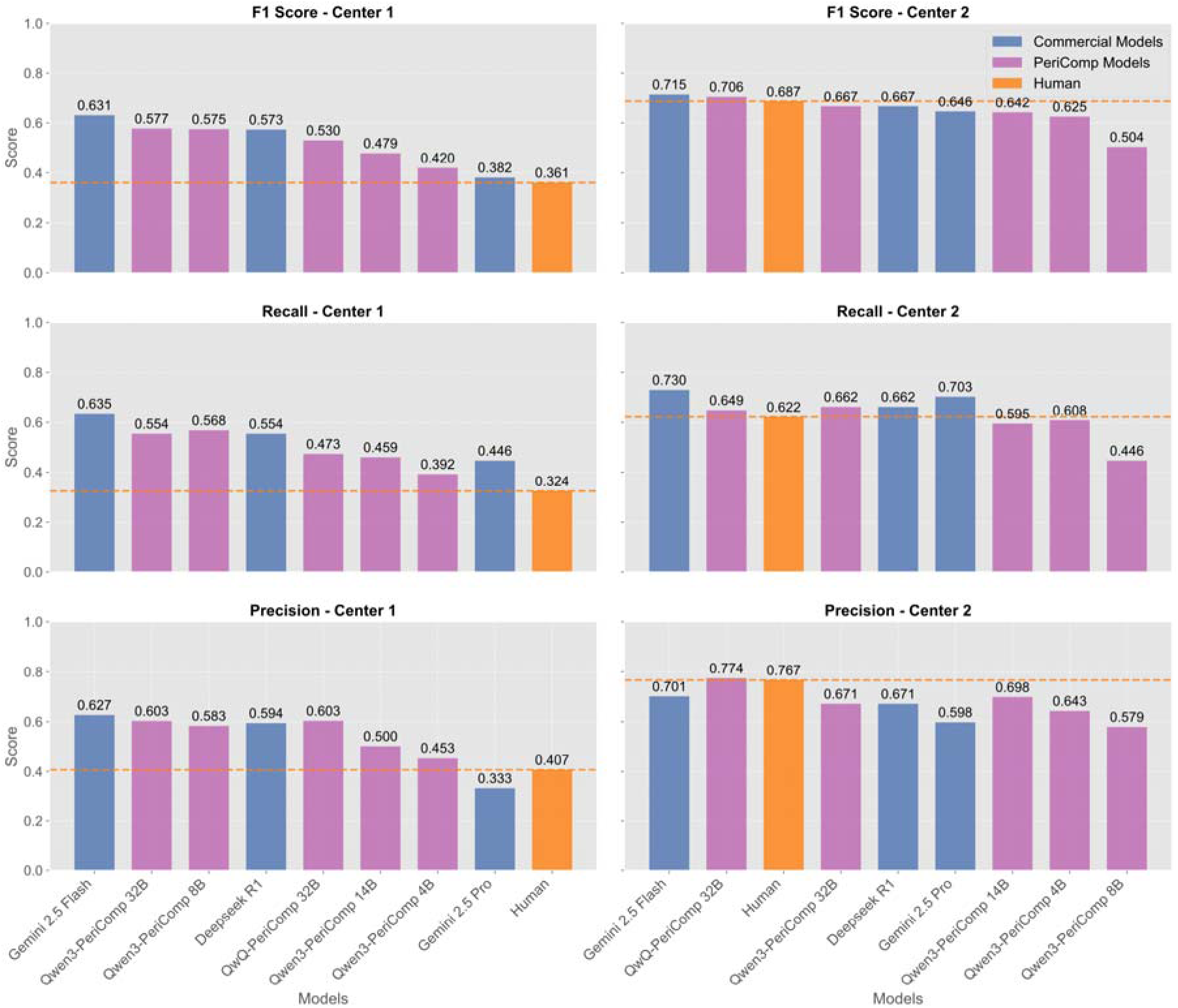
Strict performance evaluation with complete diagnostic accuracy requirements. Comprehensive performance comparison across F1 score, recall, and precision metrics for both Center 1 and Center 2 validation datasets under strict evaluation criteria, requiring correct identification of both specific complication type and severity grade (mild, moderate, or severe) for a diagnosis to be considered valid. Commercial models (blue) including Gemini 2.5 Flash, DeepSeek R1, and Gemini 2.5 Pro are compared against fine-tuned PeriComp models (purple) including QwQ-PeriComp-4B, QwQ-PeriComp-8B, QwQ-PeriComp-14B, and QwQ-PeriComp-32B, with human expert performance (orange) serving as clinical benchmarks. Orange dashed lines indicate human expert performance thresholds for each metric in respective centers. Results demonstrate that fine-tuned PeriComp models substantially approach or exceed leading commercial models and human expert performance across both validation datasets.

**Table 2.**
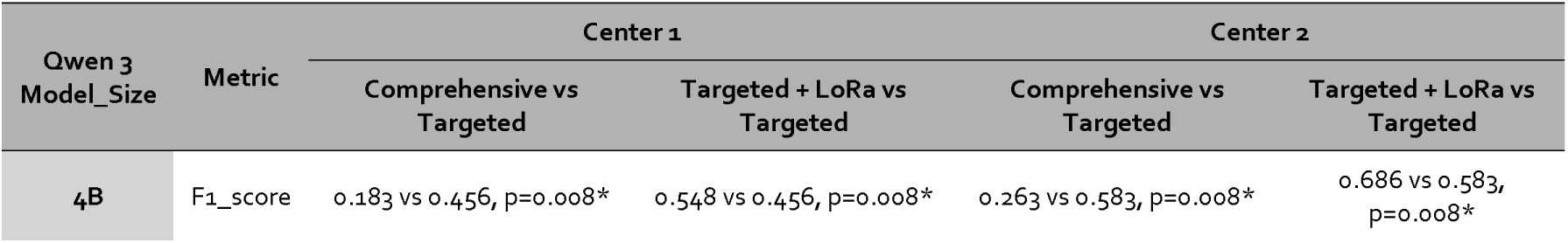
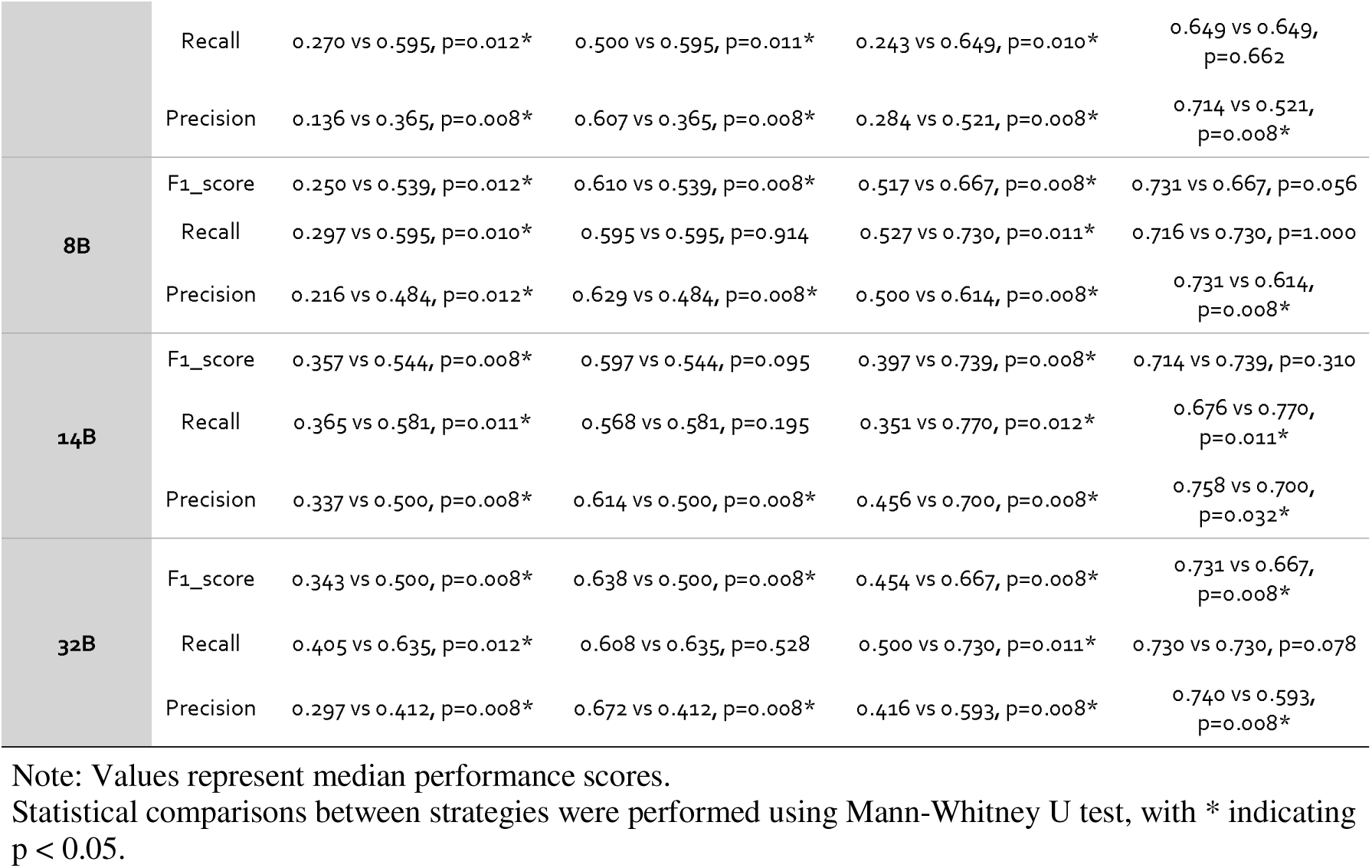
Statistical Comparison of Micro-averaged Performance Metrics Across Different Strategies and Model Sizes.

In the Center 1 dataset, the 4B model showed dramatic enhancement: its F1 score increased from approximately 0.18 to 0.45 after targeted strategy implementation and further rose to 0.55 post-LoRa fine-tuning, nearing the human performance benchmark of 0.526. The 8B model similarly demonstrated significant gains, achieving an F1 score above 0.61 after fine-tuning and surpassing human expert performance. Comparable trends were noted in the Center 2 dataset. The 14B model showed minimal improvement post-LoRa fine-tuning, suggesting its baseline performance was near its inherent capacity.

To foster research transparency and stimulate further advancements, our fine-tuned models have been made publicly available on the Hugging Face platform with the suffix “-PeriComp” appended to their original names (https://huggingface.co/collections/gscfwid/pericomp). Open-sourcing these specialized models aims to empower other researchers and drive further breakthroughs in clinical AI.

### 8. Strict performance

To further validate clinical utility of our optimized models and rigorously compare our fine-tuned models with leading commercial/cloud counterparts, we implemented a more stringent evaluation protocol. This strict assessment mandated complete diagnostic accuracy: correct identification of both the specific complication type and its severity grade (mild, moderate, or severe) was required for a diagnosis to be deemed valid.

The strict performance evaluation (Fig. 7 showing micro-averaged metrics; macro-averaged results in Appendix Fig. 8) revealed key insights into model capabilities and methodological approach efficacy. Although absolute scores were lower across all models compared to non-strict evaluations, the performance of our fine-tuned models substantially approached or exceeded that of leading commercial/cloud models and human experts across both validation datasets. Notably, human expert performance exhibited substantial variation based on case complexity: when the same experts evaluated Center 1 cases, they achieved F1 scores below 0.4, a stark contrast to their substantially higher performance on Center 2 cases, consistent with our earlier findings (Fig. 3a).

## Discussion

This study advances perioperative complication detection through several key innovations. We introduced a novel prompt framework enabling simultaneous identification and severity grading via structured output, demonstrating AI’s capacity to match or surpass human expert performance. Comprehensive evaluations identified QwQ 32B as achieving optimal balance between performance and deployment efficiency. To improve accessibility and efficacy of smaller models, we first implemented a task decomposition strategy (targeted strategy), which markedly elevated their capabilities to expert level even before fine-tuning. Subsequent LoRa fine-tuning of compact models (4B–8B parameters), leveraging this decomposed framework, further consolidated and augmented this expert-level performance, ensuring they could rival or exceed human expertise while maintaining practical deployability. Our optimized models and code have been open-sourced to foster advancements in perioperative AI.

Our task decomposition strategy aligns with established principles in machine learning, notably in modular learning and prompt engineering. Recent progress in decomposed prompting has shown substantial improvements in complex reasoning tasks by segmenting problems into manageable, sequentially solvable subtasks^19^. This modular methodology has been validated in neuro-symbolic learning frameworks, where systematic decomposition benefits complex reasoning tasks more than end-to-end learning^20^. Our clinical application of this principle reconfigures cognitive load distribution from concurrent multi-task processing to sequential single-task optimization. This shift enables sophisticated diagnostic capabilities in resource-limited models, maintaining performance comparable to that of larger, more computationally intensive alternatives.

Our strict evaluation (Fig. 7) highlighted marked heterogeneity in human expert performance between centers, with experts at Center 1 achieving F1 scores below 0.4 compared to substantially higher performance at Center 2. This inter-center disparity corroborates our earlier observations of human performance deteriorating with increasing document complexity and cognitive load, potentially owing to fatigue^17,18^. This institutional difference underscores a fundamental advantage of AI models: consistent performance across varying complexity levels, devoid of fatigue-related degradation observed in human experts processing lengthy, complex clinical records. This characteristic renders AI particularly well-suited for challenging clinical environments where comprehensive documentation and intricate case presentations are standard.

Previous research laid important foundations for AI applications in perioperative medicine. Traditional machine learning approaches demonstrated effectiveness across specific complications or narrow procedural contexts, typically yielding binary prediction outputs^21–24^. Recent LLM studies showed promise: Chung et al. (2024) examined GPT-4 Turbo’s capabilities in perioperative risk prediction using real-world electronic health records^25^, while Hsueh et al. (2024) investigated GPT-4’s capacity to detect postoperative complications from renal surgery discharge summaries^4^. However, both studies relied exclusively on commercial cloud-based models and did not explore systematic performance enhancement strategies, posing significant barriers to clinical deployment arising from data sovereignty, patient privacy, and computational cost concerns.

In contrast, our study presents a comprehensive framework addressing these limitations via iterative model enhancement. Through targeted task decomposition and LoRa fine-tuning, we systematically optimized model performance in processing unstructured clinical narratives directly. Our approach facilitates simultaneous detection and severity grading across 22 distinct perioperative complication types based on European Perioperative Clinical Outcome (EPCO) definitions—achieving complexity and coverage that surpasses prior work. More critically, our targeted strategy significantly boosts performance of smaller, resource-efficient models, enabling them to match or exceed both human expert capabilities and commercial model benchmarks while remaining deployable within existing healthcare infrastructure.

The observed performance disparities among open-source models (Fig. 4a) can be partially attributed to language-specific distributions in training data. Our exclusive use of Chinese medical records naturally favored models with substantial Chinese language representation in their pre-training corpora. For instance, Qwen models, designed with significant Chinese content (approximately 22.7% Chinese text data per Bai et al., 2023)^26^, outperformed Gemma and Mistral models, which were predominantly trained on

English-centric datasets with minimal Chinese representation. This language alignment effect is well-documented in multilingual model research, where performance correlates strongly with the quantity and quality of language-specific training data^27^. The differing performance patterns across model families likely reflect this fundamental bias in training data composition, underscoring the importance of selecting language-matched models for optimal clinical deployment in non-English healthcare settings.

Our targeted strategy combined with LoRa fine-tuning empowers 4B–8B parameter models to achieve expert-level performance, rendering perioperative complication detection an economically viable clinical tool. This advancement enables healthcare institutions to implement real-time automated screening systems providing continuous 24/7 “second opinions” to clinical teams, potentially dramatically reducing the 27% under-reporting rate associated with manual detection methods^6^. The capacity to operate within existing infrastructure while preserving data sovereignty makes this approach especially valuable for resource-limited settings.

Several limitations warrant acknowledgment. First, the exclusive use of Chinese-language clinical records raises questions regarding cross-linguistic generalizability, although the underlying targeted strategy and LoRa fine-tuning methodology are expected to be broadly applicable. Second, our approach is fundamentally reliant on the accuracy and completeness of clinical documentation by surgical teams; despite achieving expert-level performance, the models are inherently constrained by input data quality, where incomplete symptom records, absent examination findings, or inaccurate postoperative observations could precipitate diagnostic errors. Third, our training cohort, despite clinical diversity, did not encompass the complete spectrum of surgical procedures and complication patterns in contemporary perioperative practice. Future research incorporating more comprehensive and varied datasets—spanning additional surgical specialties, rare complications, and diverse institutional practices—could potentially yield further performance enhancements and bolster the robustness of our diagnostic framework.

In conclusion, this study signifies notable advancement in perioperative complication detection by establishing a comprehensive framework that marries cutting-edge AI capabilities with practical clinical deployment constraints. Our innovative amalgamation of targeted strategy and LoRa fine-tuning democratizes expert-level diagnostic performance, enabling resource-limited healthcare institutions to deploy sophisticated AI systems using modest computational infrastructure. Through open-sourcing our fine-tuned models and comprehensive prompt templates, supported by rigorous dual-center validation, we offer a foundational platform for future research and clinical implementation, providing a blueprint for developing specialized medical AI systems that balance performance, accessibility, and deployment considerations.

## Methodology

### Study Design and Data Collection

This study utilized a dual-center design to assess LLM performance in detecting postoperative complications from clinical narratives. Conducted in adherence to ethical principles, the research received approval from institutional review boards of both participating medical centers (First Affiliated Hospital of Sun Yat-sen University Ethics Committee Approval: No. [2025] 019; Jiangmen Central Hospital Ethics Committee Approval: No. [2025] 180A). The dual-center approach was chosen to evaluate model generalizability across varied healthcare environments and patient populations with distinct complexity profiles.

#### Institutional Characteristics

Center 1, a regional tertiary medical center, is characterized by complex cases, extensive clinical documentation, and a broad spectrum of postoperative complications. It serves as a referral hub for challenging surgical cases, which typically involve longer clinical narratives and more diverse complication patterns. In contrast, Center 2 is a secondary-level hospital exhibiting characteristics typical of community healthcare settings, such as relatively straightforward cases and concise documentation.

#### Inclusion and Exclusion Criteria

Inclusion criteria were formulated to capture a representative sample of complex surgical cases with adequate postoperative observation periods. Specifically, eligible patients were aged >14 years; for Center 1, an additional criterion mandated postoperative hospital stays exceeding 7 days to ensure appropriate case complexity. No restrictions were imposed on surgical specialty or procedure type.

#### Data Extraction Protocol

Electronic health records from the two independent medical centers, covering January 2023 to February 2025, were systematically extracted. For every selected case, four core data components were systematically extracted: (1) demographic and procedural information, (2) postoperative medical records, including daily clinical notes and nursing documentation, (3) abnormal postoperative laboratory results, and (4) postoperative imaging and examination reports in narrative text format.

Following systematic extraction and exclusion criteria application, the final cohort consisted of 146 cases from Center 1 and 52 cases from Center 2, totaling 198 cases for subsequent model evaluation and enhancement.

#### Data Partitioning Strategy

A portion of Center 1 data was strategically utilized for model fine-tuning, with the remainder of Center 1 cases and all Center 2 cases reserved for validation. Specifically, the 146 cases from Center 1 were randomly divided into a training cohort (n=100) for fine-tuning and a validation cohort (n=46) for performance assessment. All 52 cases from Center 2 were allocated for external validation to evaluate model generalizability. Given the heterogeneous characteristics between the two centers, we used only Center 1 data for comprehensive model comparisons and fine-tuning processes, while Center 2 data was reserved exclusively for external validation to ensure robust assessment of model generalizability.

### Data Preprocessing and Anonymization

All clinical narratives were standardized into a consistent markdown format to ensure uniform prompt integration. This structured methodology organized information hierarchically, encompassing general patient information, chronological postoperative medical records, abnormal laboratory findings, and examination reports.

Comprehensive data anonymization was performed to safeguard patient privacy. All direct identifiers were systematically removed. For temporal data, a sophisticated time-shifting methodology was implemented. We applied a unique, randomly generated temporal offset (in days) to each patient’s time-related data. This offset, inspired by established practices in medical databases like MIMIC^28^, was uniformly applied to all fields, effectively preserving the relative temporal relationships and intervals between clinical events.

### Prompt Engineering Framework

#### Role Definition and Task Specification

The prompt architecture designated the LLM as an experienced surgeon with specialized expertise in perioperative medicine, with objective explicitly defined as systematic identification and severity grading of postoperative complications based on clinical documentation and standardized diagnostic criteria.

#### Diagnostic Standards Selection

The EPCO definitions framework was selected over alternative systems, such as the Clavien-Dindo classification^29,30^. This framework delineates 22 distinct perioperative complications—most stratified into mild, moderate, and severe categories—providing comprehensive coverage with specific diagnostic criteria for each complication type.

#### Output Structure and Format Specification

A rigorous JSON schema-based output format, consistent with current OpenAI Software Development Kit standards, was implemented (as exemplified in Fig. 2c & 2e). Each complication was structured as an object with two primary attributes: complication type and severity grade. All identified complications were organized into a standardized list, with “no postoperative complications” serving as the single-element response if no complications were detected.

#### CoT Integration

As a subsequent optimization strategy, we evaluated CoT prompting incorporation to enhance diagnostic transparency and reasoning validation. When implemented, CoT-enhanced prompts included a “think” object containing structured reasoning elements.

A complete example of the structured prompt template is available in Appendix for reference and reproducibility.

### Model Selection and Deployment Architecture

The evaluation encompassed state-of-the-art commercial models including general and reasoning models from OpenAI, Anthropic, Google, Grok, and DeepSeek^31,32^. Open-source models evaluated included DeepSeek R1 & V3, QwQ 32B, the Qwen 3 series (4B, 8B, 14B, 32B), Gemma 3 (27B), and Mistral 3 Small 24B^33,34^. Detailed version specifications for all models are provided in Appendix Table 2.

Commercial models and large open-source models were accessed via established Application Programming Interface (API) services. Smaller open-source models were deployed locally on computational nodes at the National Supercomputer Center in Guangzhou, using high-performance A800 (80GB) GPU clusters with 1–4 GPUs employed in parallel configuration.

We employed MS-Swift as our fine-tuning framework for LoRa implementation^35^. Key LoRa parameters were optimized: lora_rank=16, lora_alpha=32, and learning_rate=1e-4. Local model deployment and inference utilized vLLM with integrated Outlines support for structured output generation^36,37^.

### Inference Parameters and Reproducibility

Given the logical reasoning nature of complication detection, we initially considered conservative parameters (temperature=0). However, empirical evaluation revealed that even with deterministic settings and fixed seeds, complete reproducibility remained elusive, with some models exhibiting infinite loop behaviors. Consequently, we adopted balanced parameters (temperature=0.6, top_p=0.95) that maintained reasonable determinism while preventing pathological behaviors. For commercial models, single inference calls were performed to balance robustness with cost efficiency. Local deployments utilized five repeated inferences per case to enhance statistical reliability and account for inherent stochasticity.

### Human Expert Annotation and Gold Standard Development

Human expert evaluation utilized a rigorous, blinded assessment protocol. Identical prompts to those provided to the LLMs were presented to three experienced attending physicians specializing in perioperative medicine. Cases were randomly distributed among these three experts to ensure unbiased evaluation.

The development of our gold standard incorporated both human expert opinions and initial LLM predictions as comprehensive reference points. All diagnostic outputs underwent systematic review by the three-physician panel. Final gold standard determinations necessitated a majority consensus (≥2 physicians). This gold standard served dual purposes: model evaluation and training data for fine-tuning. For fine-tuning of reasoning models, specifically QwQ 32B, all experts meticulously reviewed and refined the model’s reasoning outputs. The revised reasoning subsequently served as ‘golden reasoning’ templates, enclosed within tags, during the fine-tuning process.

### Evaluation Metrics and Statistical Analysis

We employed standard multi-class classification metrics including F1-score, precision, and recall, calculated using both micro-averaged and macro-averaged approaches. F1-score served as the primary performance indicator due to its balanced consideration of both precision and recall, particularly important in clinical contexts.

Micro-averaged metrics were calculated by aggregating true positives, false positives, and false negatives across all cases globally before computing final metrics. Macro-averaged metrics were computed by calculating precision, recall, and F1-score for each individual case and then averaging these scores across all cases. Specific computational formulations are detailed in the Appendix methodology section.

All data preprocessing, model evaluation, and statistical analyses were conducted using Python within Jupyter notebook environments. The complete codebase is slated for open-source release (https://github.com/gscfwid/PeriComp), accompanied by representative data examples. The full dataset will be accessible to qualified researchers upon reasonable request and with appropriate institutional review board approval.

## Ethics Statement

The research received approval from institutional review boards of both participating medical centers (First Affiliated Hospital of Sun Yat-sen University Ethics Committee Approval: No. [2025] 019; Jiangmen Central Hospital Ethics Committee Approval: No. [2025] 180A).

## Data Availability

Clinical datasets can be requested from corresponding authors with appropriate institutional approval.

