## Appendix for "Enhancing Privacy-Preserving Deployable Large Language Models for Perioperative Complication Detection: A Targeted Strategy with LoRA Fine-tuning"

### Appendix Figures

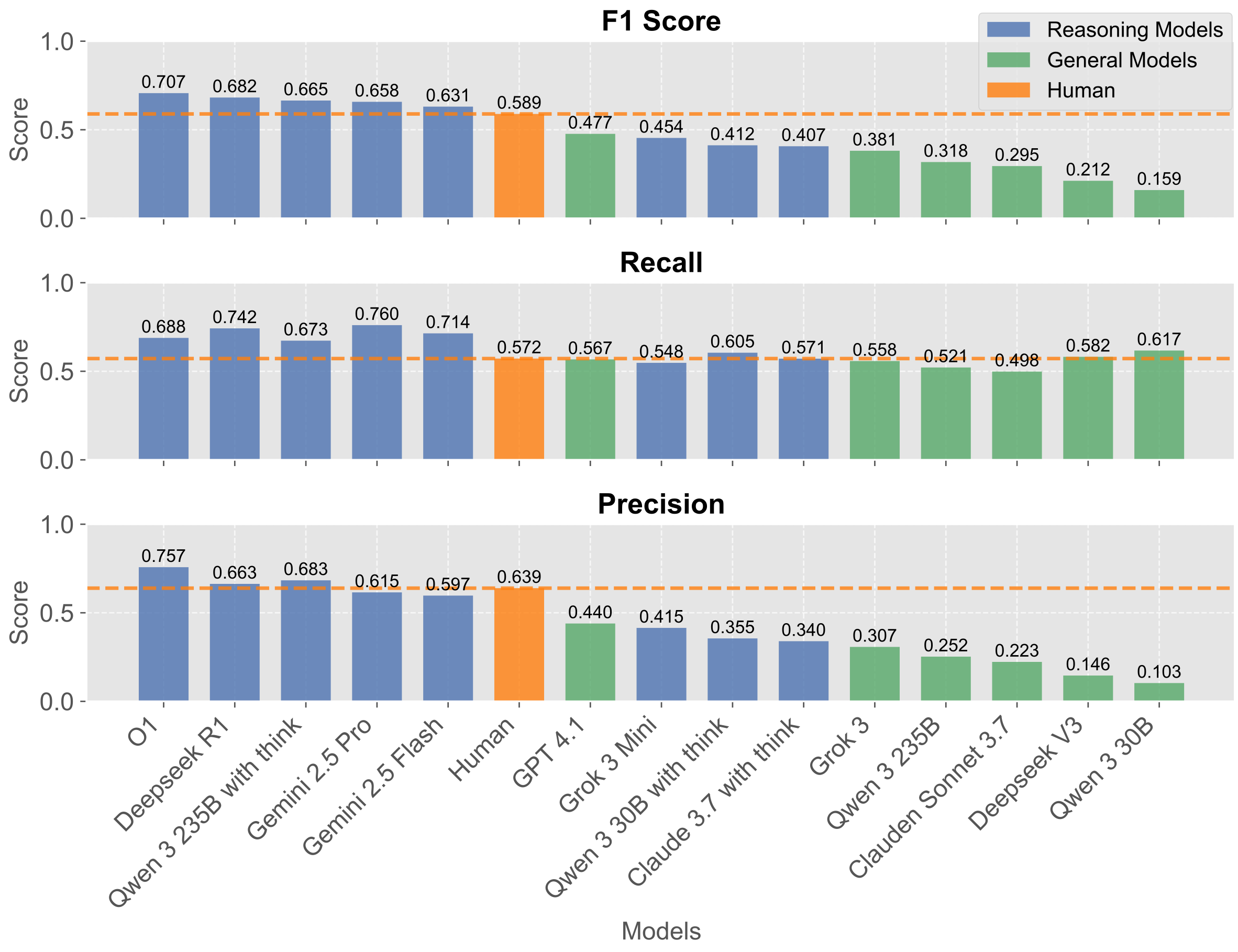

****Appendix Figure 1. Initial model performance evaluation using macro-averaged metrics.**** Performance assessment of multiple language models using basic structured JSON output format. Macro-averaged F1 score, recall, and precision metrics for reasoning models (blue), general models (green), and human experts (orange). Orange dashed lines indicate human expert benchmarks. This complements the micro-averaged results in Figure 2d.

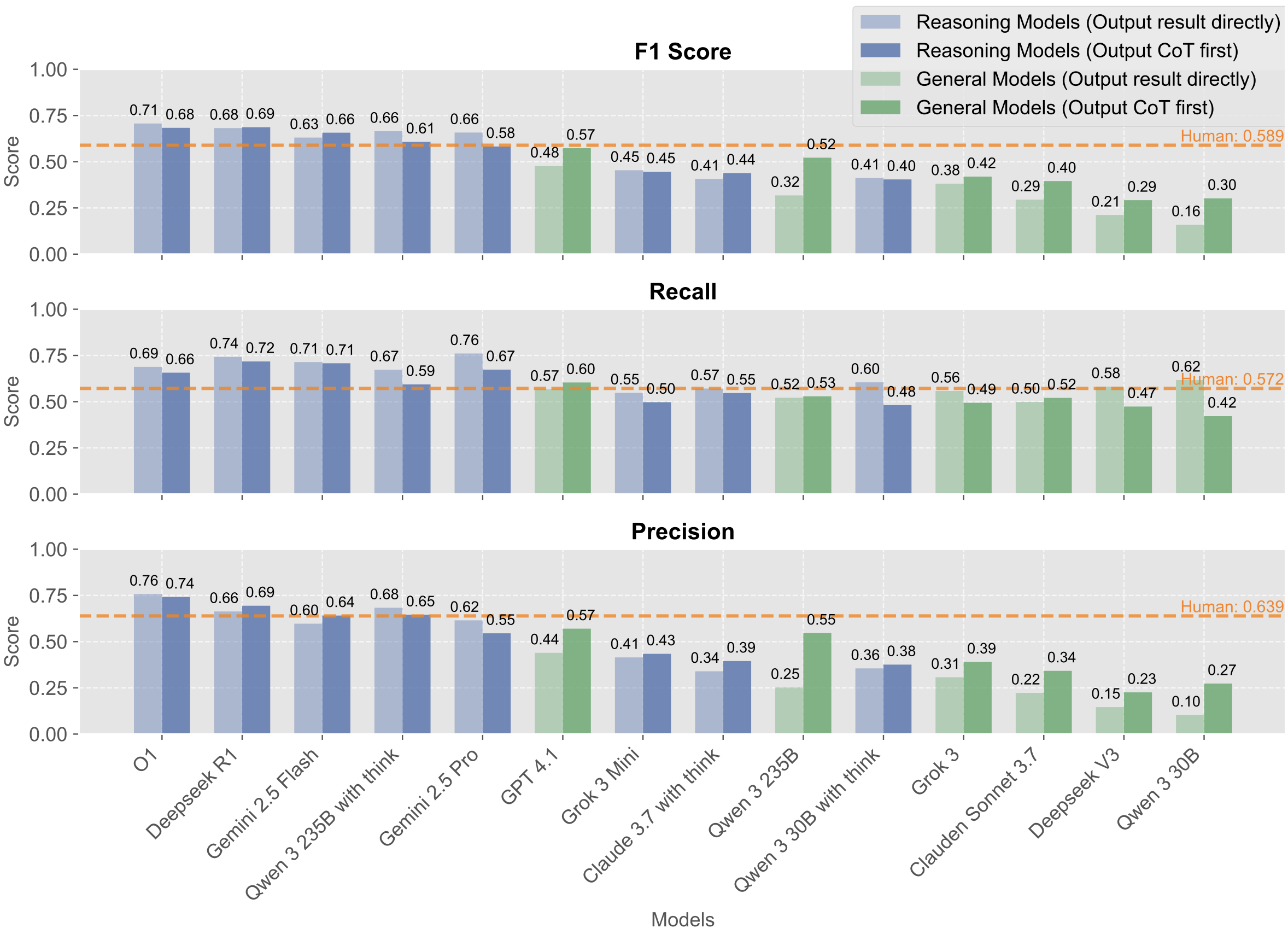

****Appendix Figure 2. Chain-of-Thought prompting performance comparison using macro-averaged metrics.**** Performance comparison before and after Chain-of-Thought (CoT) implementation across multiple language models. Macro-averaged F1 score, recall, and precision metrics showing reasoning models with direct output (light blue) versus CoT output (dark blue), and general models with direct output (light green) versus CoT output (dark green). Orange dashed lines indicate human expert benchmarks. This complements the micro-averaged results in Figure 2f.

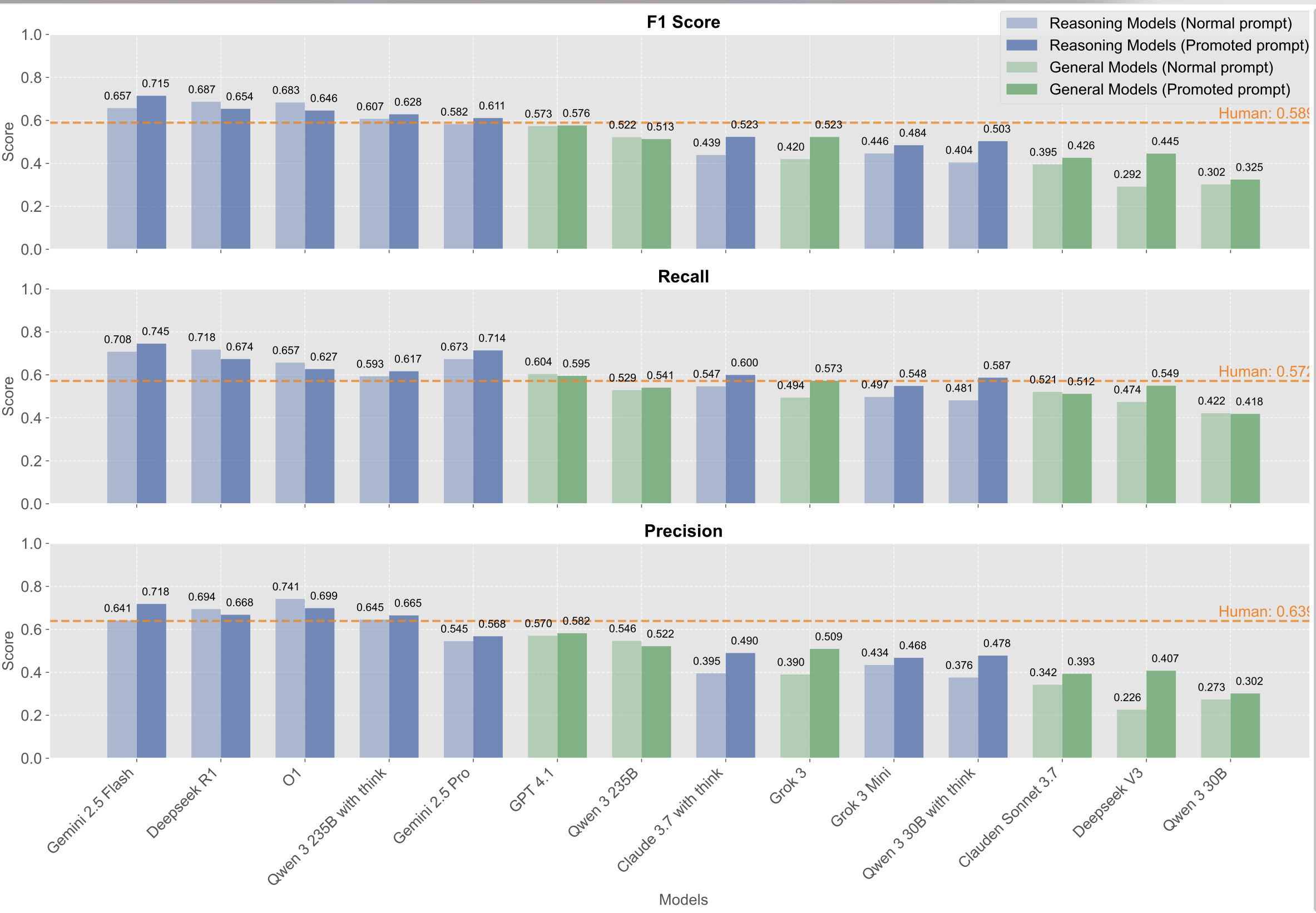

****Appendix Figure 3. Prompt optimization performance comparison using macro-averaged metrics.**** Performance comparison before and after prompt optimization across multiple language models. Macro-averaged F1 score, recall, and precision metrics showing reasoning models with normal prompts (light blue) versus promoted prompts (dark blue), and general models with normal prompts (light green) versus promoted prompts (dark green). Orange dashed lines indicate human expert benchmarks. Results demonstrate substantial improvements in initially lower-performing models following systematic prompt refinements. This complements the micro-averaged results in Figure 3c.

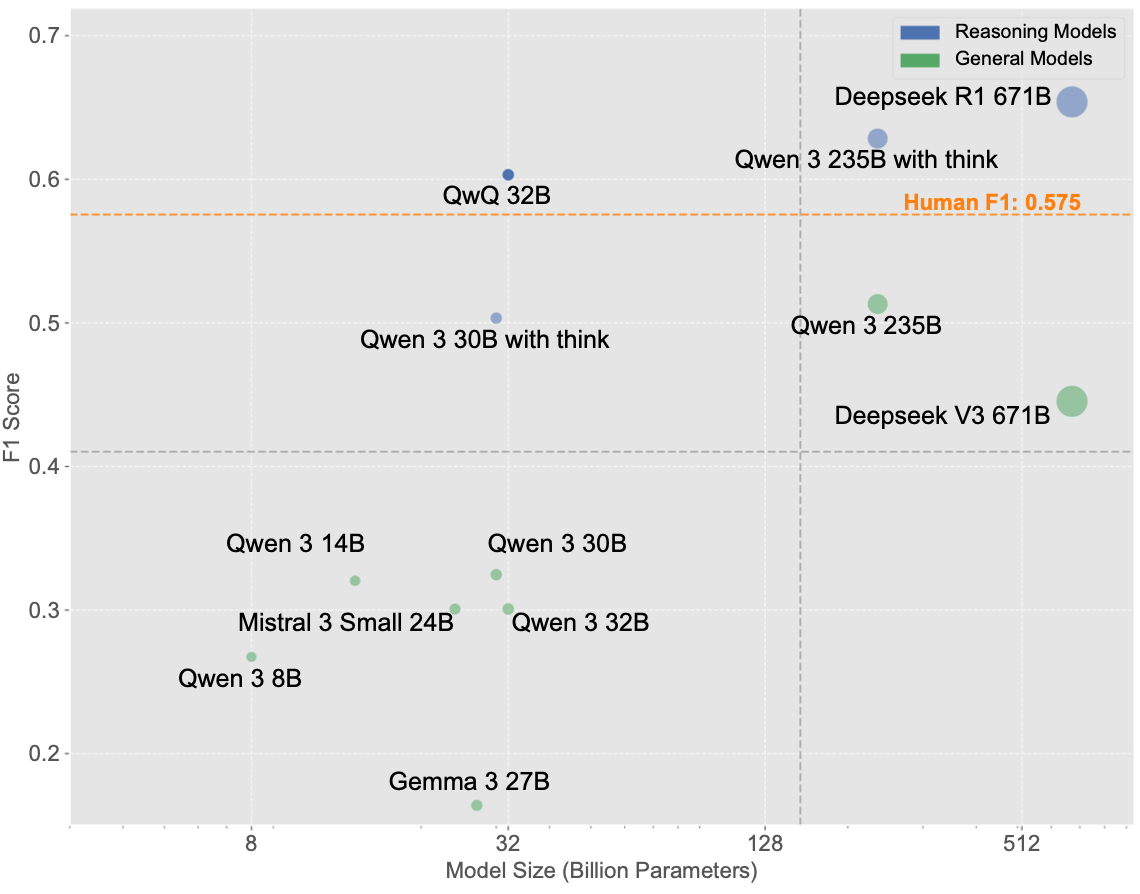

****Appendix Figure 4. Open-source model performance quadrant analysis using macro-averaged metrics.**** Quadrant chart showing systematic evaluation of open-source models spanning 4B to 671B parameters with macro-averaged F1 scores plotted against model size. Point sizes are proportional to model parameter counts. Reasoning models (blue) and general models (green) demonstrate correlation between model size and performance. Orange dashed line indicates human expert benchmark (F1 = 0.575). This complements the micro-averaged results in Figure 4a.

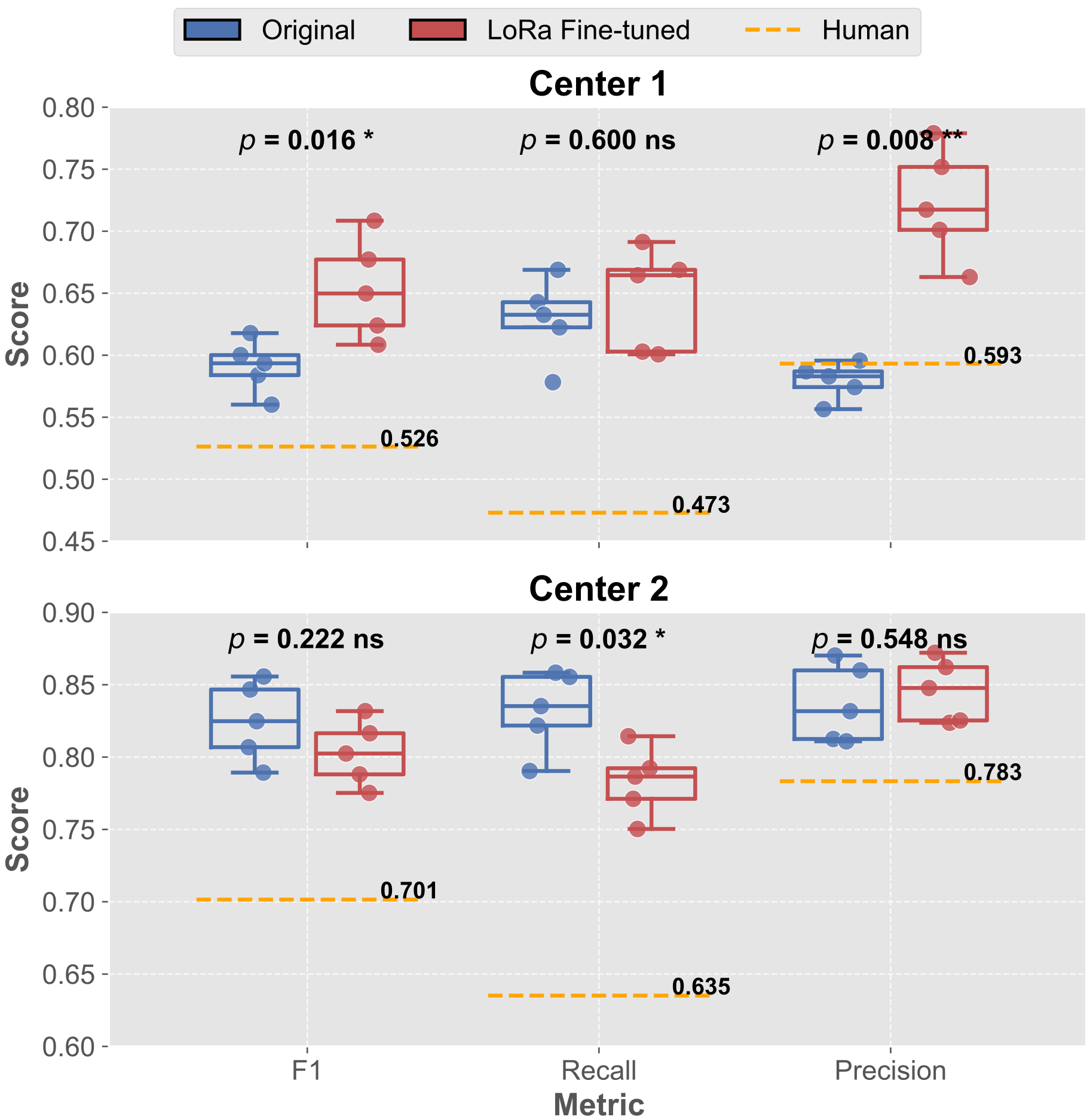

****Appendix Figure 5. QwQ 32B LoRa fine-tuning performance evaluation using macro-averaged metrics.**** Box plot comparison showing performance before (blue) and after (red) LoRa fine-tuning for QwQ 32B model across F1 score, recall, and precision metrics on both Center 1 and Center 2 datasets. Orange dashed lines indicate human expert benchmarks for each center. Statistical significance testing results are shown with p-values (* p < 0.05, ** p < 0.01, ns = not significant). This complements the micro-averaged results in Figure 4c.

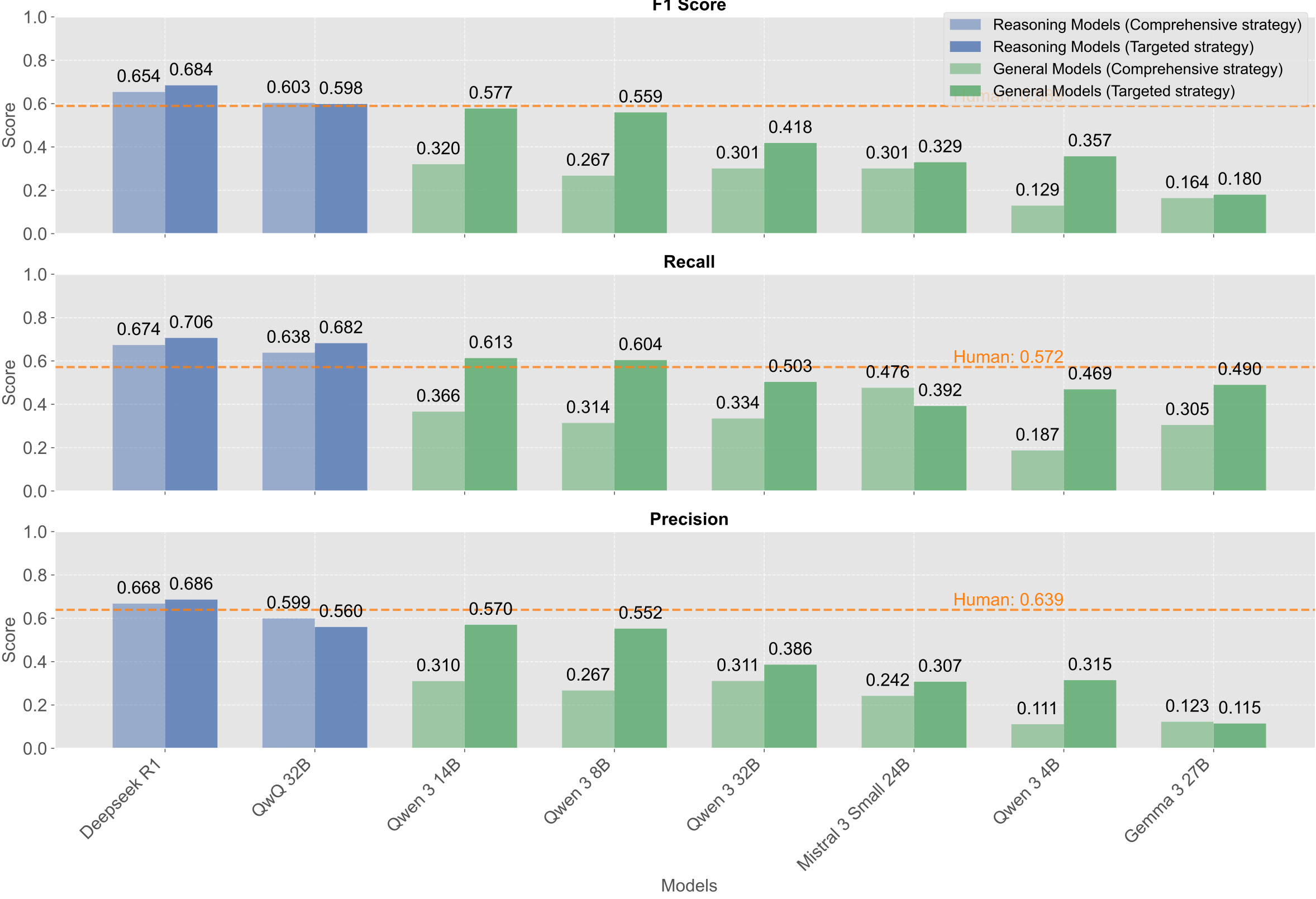

****Appendix Figure 6. Comprehensive versus targeted strategy comparison using macro-averaged metrics.**** Performance comparison between comprehensive strategy (lighter colors) and targeted strategy (darker colors) across multiple models. Macro-averaged F1 score, recall, and precision metrics for reasoning models (light blue vs. dark blue) and general models (light green vs. dark green). Orange dashed lines indicate human expert benchmarks. Results demonstrate that smaller models benefit substantially from the targeted approach while larger reasoning models maintain consistent performance. This complements the micro-averaged results in Figure 5b.

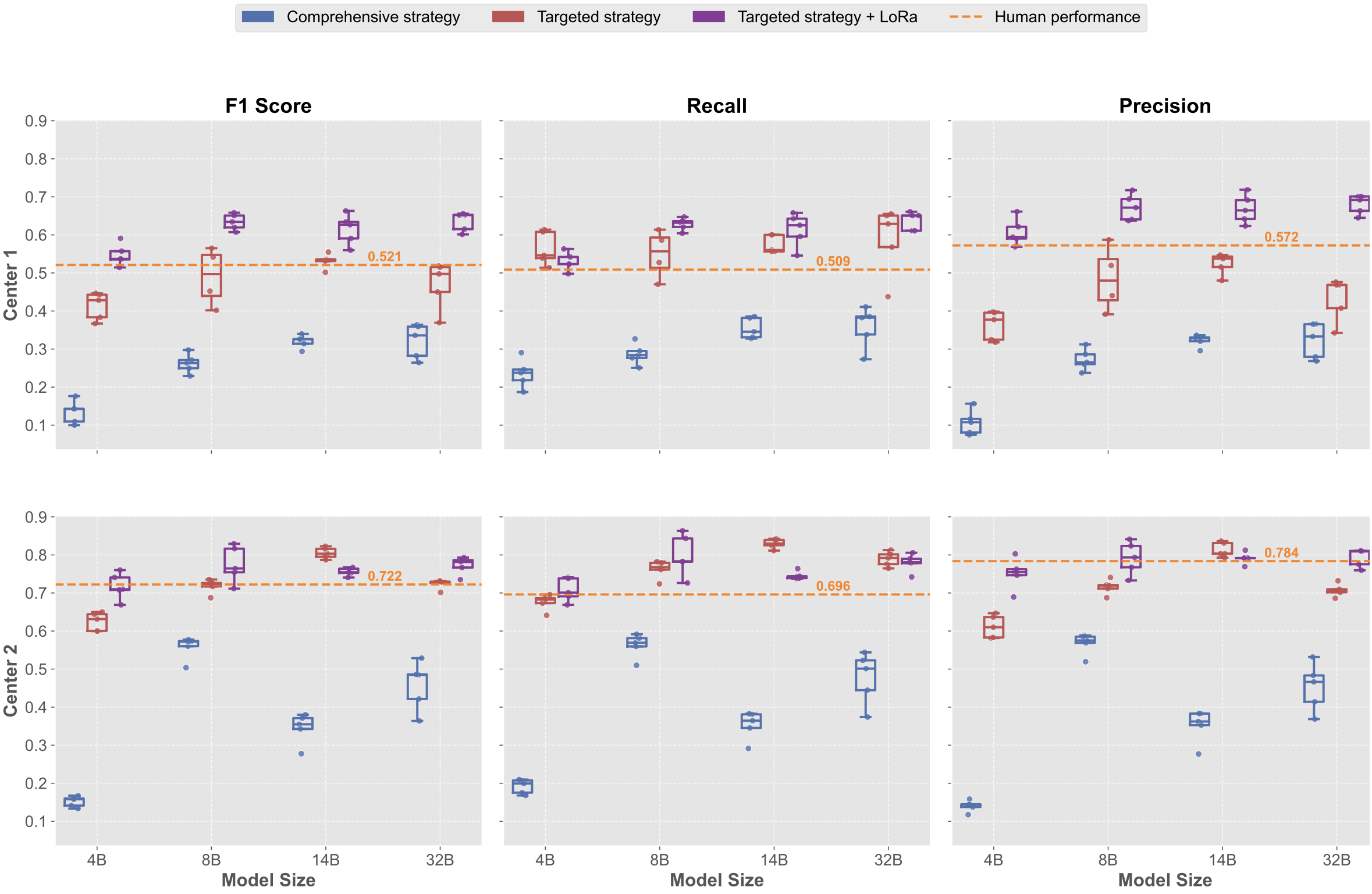

****Appendix Figure 7. LoRa fine-tuning optimization results for Qwen 3 models using macro-averaged metrics.**** Comprehensive performance comparison across F1 score, recall, and precision metrics for Qwen 3 models (4B, 8B, 14B, 32B parameters) across comprehensive strategy (blue), targeted strategy (red), and targeted strategy with LoRa fine-tuning (purple) on both Center 1 and Center 2 validation datasets. Box plots show performance distributions with orange dashed lines indicating human expert benchmarks. Results demonstrate significant performance improvements in smaller models (4B and 8B) following targeted strategy plus LoRa fine-tuning. This complements the micro-averaged results in Figure 6b.

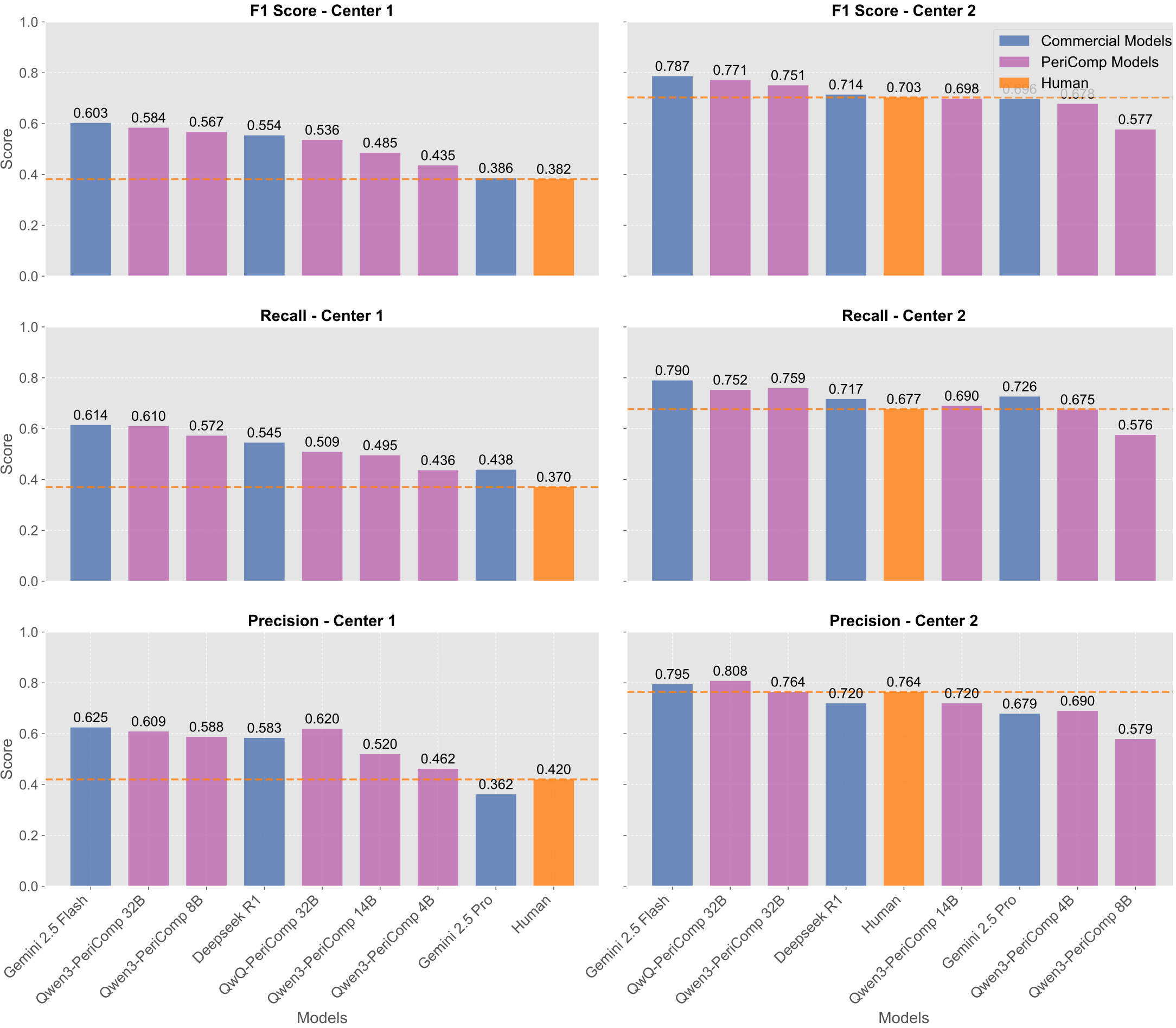

****Appendix Figure 8. Strict performance evaluation with PeriComp models using macro-averaged metrics.**** Performance comparison under strict evaluation criteria requiring correct identification of both complication type and severity grade. Macro-averaged F1 score, recall, and precision metrics comparing commercial models (blue) including Gemini 2.5 Flash and DeepSeek R1, fine-tuned PeriComp models (purple) including QwQ-PeriComp series (32B, 8B, 14B, 4B), and human expert performance (orange) across Center 1 and Center 2 datasets. Orange dashed lines indicate human expert benchmarks for each center and metric. This complements the micro-averaged results in Figure 7.

### Appendix Tables

| **Table 1. Statistical Comparison of Macro-averaged Performance Metrics Across Different Strategies and Model Sizes** | | | | | |
| --- | --- | --- | --- | --- | --- |
| **Model_Size** | **Metric** | **Center 1** | | **Center 2** | |
|  |  | **Comprehensive vs Targeted** | **Targeted + LoRa vs Targeted** | **Comprehensive vs Targeted** | **Targeted + LoRa vs Targeted** |
| **4B** | F1_score | 0.142 vs 0.428, p=0.008* | 0.537 vs 0.428, p=0.008* | 0.158 vs 0.631, p=0.008* | 0.710 vs 0.631, p=0.008* |
|  | Recall | 0.238 vs 0.546, p=0.008* | 0.523 vs 0.546, p=0.295 | 0.199 vs 0.683, p=0.008* | 0.701 vs 0.683, p=0.151 |
|  | Precision | 0.108 vs 0.377, p=0.008* | 0.594 vs 0.377, p=0.008* | 0.144 vs 0.610, p=0.008* | 0.755 vs 0.610, p=0.008* |
| **8B** | F1_score | 0.263 vs 0.529, p=0.008* | 0.634 vs 0.529, p=0.008* | 0.572 vs 0.724, p=0.008* | 0.765 vs 0.724, p=0.095 |
|  | Recall | 0.284 vs 0.586, p=0.008* | 0.632 vs 0.586, p=0.016* | 0.569 vs 0.768, p=0.008* | 0.783 vs 0.768, p=0.116 |
|  | Precision | 0.265 vs 0.520, p=0.008* | 0.672 vs 0.520, p=0.008* | 0.575 vs 0.720, p=0.008* | 0.794 vs 0.720, p=0.016* |
| **14B** | F1_score | 0.326 vs 0.533, p=0.008* | 0.627 vs 0.533, p=0.008* | 0.355 vs 0.803, p=0.008* | 0.755 vs 0.803, p=0.008* |
|  | Recall | 0.346 vs 0.560, p=0.012* | 0.625 vs 0.560, p=0.295 | 0.364 vs 0.830, p=0.008* | 0.743 vs 0.830, p=0.008* |
|  | Precision | 0.328 vs 0.538, p=0.008* | 0.664 vs 0.538, p=0.008* | 0.362 vs 0.803, p=0.008* | 0.792 vs 0.803, p=0.059 |
| **32B** | F1_score | 0.336 vs 0.497, p=0.008* | 0.652 vs 0.497, p=0.008* | 0.485 vs 0.729, p=0.008* | 0.782 vs 0.729, p=0.008* |
|  | Recall | 0.383 vs 0.629, p=0.008* | 0.651 vs 0.629, p=0.525 | 0.501 vs 0.792, p=0.008* | 0.781 vs 0.792, p=0.690 |
|  | Precision | 0.333 vs 0.468, p=0.032* | 0.692 vs 0.468, p=0.012* | 0.466 vs 0.706, p=0.008* | 0.809 vs 0.706, p=0.008* |

| **Table 2. Detailed Model Version Specifications** | | | | | | |
| --- | --- | --- | --- | --- | --- | --- |
| **Model Category** | **Model Name** | **Version/API** | **Parameters** | **Model Type** | **Access Method** | **Deployment Platform** |
| **Commercial Models** |  |  |  |  |  |  |
| OpenAI | GPT-4.1 | gpt-4.1-2025-04-14 | Unknown | General | API | Openrouter |
| OpenAI | O1 | o1-2024-12-17 | Unknown | Reasoning | API | Openrouter |
| Anthropic | Claude-3.7-Sonnet | claude-3-7-sonnet-20241022 | Unknown | General | API | Anthropic Console |
| Anthropic | Claude-3.7-Sonnet with think | claude-3-7-sonnet-20250219 | Unknown | Reasoning | API | Anthropic Console |
| Google | Gemini-2.5-Pro | gemini-2.5-pro-exp-03-25 | Unknown | Reasoning l | API | Google AI Studio |
| Google | Gemini-2.5-Flash | gemini-2.5-flash-preview-04-17 | Unknown | Reasoning | API | Google AI Studio |
| Grok | Grok-3 | grok-3-beta | Unknown | General | API | xAI Platform |
| Grok | Grok-3-mini | Grok-3-mini-beta | Unknown | Reasoning | API | xAI Platform |
| **Open-source Models** |  |  |  |  |  |  |
| DeepSeek | DeepSeek-V3 | deepseek-chat | 671B | General | API | DeepSeek Platform |
| DeepSeek | DeepSeek-R1 | deepseek-reasoner | 671B | Reasoning | API | DeepSeek Platform |
| Alibaba | Qwen3-235B | Qwen3 235B A22B | 235B | Reasoning | API | Openrouter |
| Alibaba | Qwen3-30B | Qwen3 30B A3B | 30B | Reasoning | API | Openrouter |
| Alibaba | QwQ-32B | QwQ-32B | 32B | Reasoning | Local | Hugging Face |
| Alibaba | Qwen3-4B | Qwen3-4B | 4B | General | Local | Hugging Face |
| Alibaba | Qwen3-8B | Qwen3-8B | 8B | General | Local | Hugging Face |
| Alibaba | Qwen3-14B | Qwen3-14B | 14B | General | Local | Hugging Face |
| Alibaba | Qwen3-32B | Qwen3-32B | 32B | General | Local | Hugging Face |
| Google | Gemma-3-27B | Gemma-3-27b-it | 27B | General | Local | Hugging Face |
| Mistral AI | Mistral-Small-3-24B | Mistral-Small-24B-Instruct-2501 | 24B | General | Local | Hugging Face |
| **Fine-tuned Models (This Study)** |  |  |  |  |  |  |
| Alibaba | Qwen3-4B-PeriComp | Qwen3-4B-PeriComp | 4B | General | Local | Hugging Face |
| Alibaba | Qwen3-8B-PeriComp | Qwen3-8B -PeriComp | 8B | General | Local | Hugging Face |
| Alibaba | Qwen3-14B- PeriComp | Qwen3-14B-PeriComp | 14B | General | Local | Hugging Face |
| Alibaba | Qwen3-32B- PeriComp | Qwen3-32B-PeriComp | 32B | General | Local | Hugging Face |
| Alibaba | QwQ-32B- PeriComp | QwQ-32B-PeriComp | 32B | Reasoning | Local | Hugging Face |
| Note: All models without specific version numbers represent the latest versions available before April 2025 | | | | | | |

### An example of prompt (Translated to English)

#### Comprehensive original version

You are a senior surgeon. Your task is to identify postoperative complications based on patient data, and the diagnostic criteria for complications are:

1 Acute Kidney Injury

- Definition: Within seven days postoperatively, meeting KDIGO criteria:

- Grade I: Creatinine 1.5–1.9 times baseline or urine output reduction for 6–12 hours

- Grade II: Creatinine 2–2.9 times baseline or urine output reduction >12 hours

- Grade III: Creatinine ≥3 times baseline or requiring renal replacement therapy

2 Acute Respiratory Distress Syndrome (ARDS)

- Bilateral infiltrates within one week of known clinical insult or new/worsening respiratory symptoms, not fully explained by effusions, lobar/lung collapse, or nodules

- Respiratory failure not fully explained by cardiac failure or fluid overload. If no risk factors present, objective assessment (such as echocardiography) is needed to rule out pulmonary edema

- Oxygenation levels:

- Mild: PaO2/FiO2 26.7-40.0 kPa (200-300 mmHg)

- Moderate: PaO2/FiO2 13.3-26.6 kPa (100-200 mmHg)

- Severe: PaO2/FiO2 ≤13.3 kPa (100 mmHg)

3 Anastomotic breakdown

- Definition: Leakage of contents from surgical connection sites, including gastrointestinal, biliary, pancreatic ducts, etc., which may lead to fever, abscess, or organ failure

- Grading:

- Mild: Asymptomatic, only imaging findings

- Moderate: Requires treatment, no permanent damage

- Severe: Requires surgical intervention or causes organ dysfunction

4 Arrhythmia

- ECG-confirmed cardiac rhythm abnormalities

- Grading: Standard grading

5 Cardiac arrest

- Cessation of mechanical cardiac activity with no circulatory signs

- Diagnostic criteria: ECG or clinical examination

- No grading

6 Cardiogenic pulmonary oedema

- Alveolar fluid accumulation due to cardiac dysfunction

- Grading: Standard grading

7 Deep vein thrombosis (DVT)

- Newly formed thrombus in the venous system detected by ultrasound, venography, or CT imaging

- Grading: Standard grading

8 Delirium

- Each criterion met scores one point:

- Inattention

- Disorientation

- Hallucinations-delusions-psychosis

- Psychomotor agitation or retardation

- Inappropriate speech or mood

- Sleep/wake cycle disturbance or symptom fluctuation

- Delirium can be diagnosed when score reaches 4 points

- No grading

9 Gastrointestinal bleed

- Clear clinical or endoscopic evidence of gastrointestinal bleeding

- Grading: Standard grading

10 Infection, source uncertain

- Clinically suspected infection with undetermined source

- Meeting two or more of the following:

- Core temperature < 36°C or > 38°C

- White blood cell count > 12 × 10^9 L^-1 or < 4 × 10^9 L^-1

- Respiratory rate >20 breaths per minute or PaCO2 < 4.7 kPa (35 mmHg)

- Heart rate >90 beats per minute

- Grading: Standard grading

11 Laboratory confirmed bloodstream infection

- At least one of the following, unrelated to infection at other sites:

- Positive blood culture with exclusion of other infection sources

- Clinical symptoms (fever, chills, or hypotension) plus supportive indicators (at least one of the following):

- a. Common skin contaminants isolated from two or more blood cultures drawn at different times

- b. Common skin contaminants isolated from intravascular catheter blood culture with appropriate antimicrobial therapy initiated by physician

- c. Positive blood antigen test

- Grading: Standard grading

12 Myocardial infarction

- Elevated cardiac injury markers (such as TnT) with one of the following:

- Ischemic symptoms

- New ECG abnormalities (such as ST changes, left bundle branch block, or pathological Q waves)

- Regional wall motion abnormalities

- Grading: Standard grading

13 Myocardial injury after non-cardiac surgery (MINS)

- Definition: TnT ≥0.03 ng/mL without other causes (including renal dysfunction)

- No grading

14 Pneumonia

- Must meet all of the following conditions:

- Radiological criteria (requires two chest X-rays if underlying lung or heart disease, otherwise one is sufficient):

- New or progressive and persistent infiltrates

- Consolidation

- Cavitation

- Systemic manifestations (at least one):

- Unexplained fever >38°C

- White blood cell count <4×10^9/L or >12×10^9/L

- For patients ≥70 years old, unexplained mental status changes

- Respiratory-related changes (at least two):

- New purulent sputum or sputum character changes, or increased secretions/need for more suctioning

- New or worsening cough, dyspnea, tachypnea

- Rales or bronchial breath sounds

- Worsening gas exchange (hypoxemia, need for increased oxygen or ventilator support)

- Grading: Standard grading

15 Paralytic ileus

- Unable to tolerate diet or absence of flatus 3 days postoperatively

- Grading: Standard grading

16 Postoperative haemorrhage

- Blood loss requiring transfusion or surgical hemostasis within 72 hours postoperatively

- Grading: Only includes moderate and severe; surgical hemostasis required is severe

17 Pulmonary embolism (PE)

- Newly formed thrombus in the pulmonary arterial system

- Grading: Standard grading

18 Stroke

- Persistent motor, sensory, or cognitive dysfunction due to embolic or hemorrhagic cerebrovascular events

- Grading: Standard grading

19 Surgical site infection (superficial)

- Infection within 30 days postoperatively

- Limited to incision skin and subcutaneous tissue

- Meeting any of the following:

- Purulent drainage from superficial incision

- Pathogenic microorganisms cultured from aseptically obtained fluid or tissue from superficial incision

- Incision site shows any of pain/tenderness, localized swelling, redness, or heat, and surgeon deliberately opens superficial incision (may be culture positive or not cultured, but culture negative does not meet criteria)

- Surgeon or attending physician diagnoses surgical site infection

- Grading: Standard grading

20 Surgical site infection (deep)

- Occurs within 30 days postoperatively if no implant, or within 1 year if implant present

- Infection involves deep soft tissues of incision (such as fascia and muscle layers)

- Patient meets at least one of the following:

- Purulent drainage from deep incision (not from organ/space)

- Deep incision spontaneously dehisces or is deliberately opened by surgeon, and patient has fever (>38°C) or local pain/tenderness, culture positive or not cultured (culture negative does not meet this criterion)

- Abscess or other evidence of infection found on direct examination, surgery, pathology, or imaging of deep incision

- Surgeon or attending physician diagnoses surgical site infection

- Grading: Standard grading

21 Surgical site infection (organ/space)

- Occurs within 30 days postoperatively

- Infection is surgery-related and involves body parts other than skin incision, fascia, muscle layers

- Patient meets at least one of the following:

- Purulent drainage from drain placed through small incision into organ/space

- Pathogenic microorganisms cultured from aseptically obtained organ/space fluid or tissue

- Abscess or other evidence of infection confirmed by direct examination, reoperation, pathology, or imaging of organ/space

- Surgeon or attending physician diagnoses organ/space surgical site infection

- Grading: Standard grading

22 Urinary tract infection

- Following two conditions appear 24h postoperatively:

- Positive urine culture (≥10^5 CFU/ml) with no more than two types of microorganisms

- Simultaneous occurrence of at least one of the following symptoms or signs: fever (>38°C), urgency, frequency, dysuria, suprapubic tenderness, costovertebral angle pain or tenderness, with no other recognized cause

- Grading: Standard grading

**### JSON Output Format**

1. First, output a think field containing an array, where each element is a string representing thoughts and considerations about the case. For example:

- "Patient recovered well postoperatively with no obvious discomfort"

- "Elevated troponin T suggests possible myocardial injury"

2. Then, output a complications field containing an array, where each element includes:

- name: Name of the complication, must be one of the 22 predefined complications

- grading: Severity grading, can only be one of the following four options:

- "Mild" - Mild: No treatment required

- "Moderate" - Moderate: Treatment required, no long-term effects

- "Severe" - Severe: Causes organ dysfunction or death, significantly prolongs hospital stay

- "Null" - No grading

Complete format example:

```json

{

"think": [

"Patient after kidney transplant surgery, acute kidney injury diagnosis excluded according to guidelines.",

"First postoperative troponin T elevated to 0.040 ng/mL, meeting definition of myocardial injury after non-cardiac surgery."

],

"complications": [

{

"name": "Myocardial Injury After Non-cardiac Surgery",

"grading": "Null"

}

]

}

```

Notes:

- If no postoperative complications are found, write "No postoperative complications" in the complications array

- If postoperative complications are found, they must be included in the above 22 complications; custom complications are prohibited

- Standard grading follows:

- Mild: No treatment required

- Moderate: Treatment required, no long-term effects

- Severe: Causes organ dysfunction or death, significantly prolongs hospital stay

**### Medical Record Data**

**# General Information**

- Gender: Male

- Age: 60-65

**# Progress Notes**

2024-05-09 19:06 First Postoperative Progress Note

- Surgery Date: May 09, 2024

- Anesthesia: General anesthesia

- Surgical Procedure: Pancreaticoduodenectomy, retroperitoneal lymph node dissection

- Intraoperative Diagnosis: Pancreatic head cancer

- Postoperative Diagnosis: Pancreatic head cancer

- Surgical Summary: Patient underwent pancreaticoduodenectomy and retroperitoneal lymph node dissection under general anesthesia today. Procedure went smoothly with 600ml blood loss, 400ml fresh frozen plasma transfused. One drain tube placed posterior to biliary-enteric anastomosis and one anterior to pancreatic-enteric anastomosis. Patient returned to ward safely. Gross specimen sent for pathological examination.

- Postoperative Management: Temporary cardiac monitoring, oxygen therapy, NPO with fluid replacement, analgesia. Blood routine and biochemistry to be rechecked tomorrow.

- Important Postoperative Observations: Monitor patient's vital signs and drainage status.

- Recorder: /

2024-05-09 20:17 Automatic Critical Value Progress Note

- 2024-05-09 19:55:31 Emergency biochemistry panel, emergency liver enzyme panel, emergency liver function panel

- Potassium K: 2.71 mmol/L ↓★

- Received Time: 2024-05-09 20:17:39

- Received by: , Processing Time: 2024-05-09 20:17:53, Processed by: , Management: Consistent with condition, must be treated, intravenous potassium supplementation provided.

- Recorder: /

2024-05-10 07:26 Professor/Attending Physician Ward Round

Today is postoperative day 1. Patient complains of wound pain, no fever, chills, chest tightness, shortness of breath, nausea, vomiting or other discomfort. Physical examination: vital signs stable, cardiopulmonary examination shows no obvious abnormalities, abdomen soft, slight reddish drainage from wound, left abdominal drainage 10ml, right abdominal drainage 20ml. Yesterday's urine output 4150ml, intake 8300ml, gastric tube drainage 2ml.

Professor's ward round instructions: Patient complains of significant postoperative wound pain, pay attention to timely symptomatic pain relief; continue liver protection, albumin, gastric protection, nutritional support today, monitor vital signs changes and wound drainage status. Follow orders.

Recorder: /

2024-05-11 08:37 Professor/Attending Physician Ward Round

Today is postoperative day 2. Patient reports wound pain improvement compared to before, oxygen saturation 90-92% without oxygen, can rise to 98% with low-flow oxygen, no fever, chills, chest tightness, shortness of breath, nausea, vomiting or other discomfort. Physical examination: vital signs stable, cardiopulmonary examination shows no obvious abnormalities, abdomen soft, slight reddish drainage from wound, left abdominal drainage 10ml, right abdominal drainage 20ml. Yesterday's urine output 2900ml, intake 3050ml, gastric tube drainage 10ml.

Laboratory Tests:

- Emergency blood routine + high-sensitivity C-reactive protein panel: WBC 12.49x10^9/L ↑, NEUT% 0.868 ↑, Hb 108g/L ↓, PLT 192x10^9/L

- Coagulation studies: PT 13.5 seconds, APTT 29.6 seconds

- Liver and biliary metabolism panel: Na 148mmol/L ↑, CREA 120umol/L ↑, ALT 63U/L ↑, AST 62U/L ↑, ALB 28.1g/L ↓, TBIL 115.6umol/L ↑

Professor's ward round instructions: Patient's postoperative oxygen saturation is decreased, monitor for postoperative cardiopulmonary complications such as pulmonary embolism, may add low molecular weight heparin for thrombosis prevention, recheck relevant tests to exclude complications.

Recorder: /

2024-05-12 07:44 Professor/Attending Physician Ward Round

Today is postoperative day 3. Patient reports wound pain improvement compared to before, oxygen saturation about 90% without oxygen, can rise to 97% with low-flow oxygen, maximum temperature yesterday 37.5℃, no chills, chest tightness, shortness of breath, nausea, vomiting or other discomfort. Physical examination: vital signs stable, cardiopulmonary examination shows no obvious abnormalities, abdomen soft, slight reddish drainage from wound, left abdominal drainage 10ml, right abdominal drainage 10ml. Yesterday's urine output 2065ml, intake 2850ml, gastric tube drainage 10ml.

Professor's ward round instructions: Patient's postoperative gastric tube drainage is low, gastric tube can be removed today, closely monitor condition changes.

Recorder: /

2024-05-15 08:35 Professor/Attending Physician Ward Round

Ward round today, patient has no obvious discomfort, no chills, chest tightness, shortness of breath, nausea, vomiting or other discomfort. Physical examination: vital signs stable, cardiopulmonary examination shows no obvious abnormalities, abdomen soft, no obvious drainage from wound.

Professor's ward round instructions: Patient's postoperative general condition is acceptable, may start feeding and discontinue 3L bag nutritional support, continue monitoring condition changes.

Recorder: /

2024-05-18 07:41 Professor/Attending Physician Ward Round

Ward round today, patient has no obvious discomfort, no chills, chest tightness, shortness of breath, nausea, vomiting or other discomfort. Physical examination: vital signs stable, cardiopulmonary examination shows no obvious abnormalities, abdomen soft, no obvious drainage from wound.

Laboratory Tests: Pathology: (Gross specimen) Tumor infiltrates pancreatic tissue, partial duodenal wall and ampulla, tumor cells show sheet-like infiltration, some in cord-like pattern, abundant cytoplasm, some cytoplasm eosinophilic, some transparent, oval nuclei, focal necrosis, neural bundle invasion and intravascular tumor emboli visible, gastric margin, small bowel margin and bile duct margin show no cancer. Also shows lymph node metastatic cancer (2/2). Specimen 15 immunohistochemistry: Cancer tissue CK7(+), CK19(+), MUC-1(+), CDX-2(-), M-CEA partial(+), INI-1(+, no loss), Brg-1(SMARCA4)(+, no loss), SMARCA2(+, no loss), Syn(-), INSM1(-), Ki-67 about 20%(+). Combined with HE morphology and immunohistochemistry results, lesion consistent with pancreatic poorly differentiated carcinoma, considered adenocarcinoma, suggest additional immunohistochemistry (Trypsin, Chymotrypsin, Bcl-10) to assist in tumor origin analysis.

Professor's ward round instructions: Patient's pancreatic cancer diagnosis is relatively clear, note that postoperative chemotherapy needs to be arranged; patient's postoperative recovery is acceptable, if no obvious discomfort, may consider discharge.

Recorder: /

2024-05-21 08:18 Professor/Attending Physician Ward Round

Ward round today, patient has no complaints of discomfort, good spirit, sleep, appetite, no dizziness, headache, fever, chills, no precordial discomfort, no abdominal distension, abdominal pain, no urinary frequency, urgency, dysuria, no gross hematuria, normal stool and urine color. Physical examination same as before.

Attending physician's ward round instructions: Patient's postoperative recovery is good, may process discharge today. Follow orders.

Recorder: /

**# Laboratory Results**

| Test Date | Test Item | Result | Unit | High/Low | Risk Flag | Reference Range |

| 2024-05-04 | Creatinine CREA | 124 | umol/L | ↑ | | 53 - 115 |

| 2024-05-09 | White Blood Cells WBC | 11.40 | x10^9/L | ↑ | | 4.00 - 10.00 |

| 2024-05-09 | Hemoglobin Hb | 103 | g/L | ↓ | | 130 - 175 |

| 2024-05-09 | Creatinine CREA | 118 | umol/L | ↑ | | 53 - 115 |

| 2024-05-10 | Creatinine CREA | 120 | umol/L | ↑ | | 53 - 115 |

| 2024-05-10 | White Blood Cells WBC | 12.49 | x10^9/L | ↑ | | 4.00 - 10.00 |

| 2024-05-10 | Hemoglobin Hb | 108 | g/L | ↓ | | 130 - 175 |

| 2024-05-12 | White Blood Cells WBC | 11.78 | x10^9/L | ↑ | | 4.00 - 10.00 |

| 2024-05-12 | Hemoglobin Hb | 93 | g/L | ↓ | | 130 - 175 |

| 2024-05-12 | Creatinine CREA | 134 | umol/L | ↑ | | 53 - 115 |

| 2024-05-14 | Hemoglobin Hb | 101 | g/L | ↓ | | 130 - 175 |

| 2024-05-14 | Creatinine CREA | 141 | umol/L | ↑ | | 53 - 115 |

| 2024-05-14 | High-sensitivity Troponin T (TnT-T) (Luminescence) | 0.241 | ng/mL | ↑ | | 0.000 - 0.014 |

| 2024-05-15 | Creatinine CREA | 159 | umol/L | ↑ | | 53 - 115 |

| 2024-05-16 | Hemoglobin Hb | 86 | g/L | ↓ | | 130 - 175 |

| 2024-05-16 | Creatinine CREA | 147 | umol/L | ↑ | | 53 - 115 |

| 2024-05-18 | Hemoglobin Hb | 90 | g/L | ↓ | | 130 - 175 |

| 2024-05-18 | D-dimer (D-D) Test | 4.21 | mg/L FEU | ↑ | | 0.00 - 0.55 |

| 2024-05-18 | Creatinine CREA | 159 | umol/L | ↑ | | 53 - 115 |

| 2024-05-20 | White Blood Cells WBC | 10.53 | x10^9/L | ↑ | | 4.00 - 10.00 |

| 2024-05-20 | Hemoglobin Hb | 94 | g/L | ↓ | | 130 - 175 |

| 2024-05-20 | Creatinine CREA | 141 | umol/L | ↑ | | 53 - 115 |

**# Bacterial Culture**

| Test Date | Sample | Test Item | Result |

| 2024-05-11 | Drainage Fluid | Preliminary Report | Preliminary report: No bacterial growth |

**# Examination Results**

2024-05-13 Imaging Diagnosis

"Radical pancreaticoduodenectomy + retroperitoneal lymph node dissection" follow-up, compared to 2024-05-07 CT:

1. Post-Whipple changes, biliary-enteric, gastro-enteric, pancreatic-enteric anastomoses patent, peritoneal exudate in surgical area, loculated fluid collection, small amount of air, abdominal drainage tubes in place.

2. Lymphatic edema around portal vein. Mild intrahepatic bile duct dilation with small amount of air. Mild pancreatic duct dilation.

3. Multiple abnormal enhancement foci in liver, possibly hemangiomas, suggest correlation with MRI examination.

4. Scattered liver cysts.

5. Multiple bilateral adrenal nodules, possibly nodular hyperplasia, suggest clinical correlation and follow-up.

6. Multiple bilateral renal stones; multiple bilateral renal cysts.

7. Bilateral lower lobe and left lingular segmental atelectasis, bilateral pleural effusion, suggest follow-up.

8. Subpleural inflammation in right middle lobe, bilateral pulmonary emphysema.

9. Coronary artery, aorta and branch atherosclerosis, aortic valve calcification.

2024-05-14 ECG Diagnosis

- Sinus rhythm

- Complete right bundle branch block

- Left ventricular high voltage

#### Comprehensive modified version

You are a senior surgeon. Your task is to identify postoperative complications based on patient data, and the diagnostic criteria for complications are:

1 Acute Kidney Injury

- Definition: Within seven days postoperatively, meeting KDIGO criteria:

- Grade I: Creatinine 1.5–1.9 times baseline or urine output reduction for 6–12 hours

- Grade II: Creatinine 2–2.9 times baseline or urine output reduction >12 hours

- Grade III: Creatinine ≥3 times baseline or requiring renal replacement therapy

- Notes:

- If the surgery is kidney transplantation, postoperative acute kidney injury should not be diagnosed

- If there are no preoperative creatinine test results, calculate based on baseline value of 84

2 Acute Respiratory Distress Syndrome (ARDS)

- Bilateral infiltrates within one week of known clinical insult or new/worsening respiratory symptoms, not fully explained by effusions, lobar/lung collapse, or nodules

- Respiratory failure not fully explained by cardiac failure or fluid overload. If no risk factors present, objective assessment (such as echocardiography) is needed to rule out pulmonary edema

- Oxygenation levels:

- Mild: PaO2/FiO2 26.7-40.0 kPa (200-300 mmHg)

- Moderate: PaO2/FiO2 13.3-26.6 kPa (100-200 mmHg)

- Severe: PaO2/FiO2 ≤13.3 kPa (100 mmHg)

3 Anastomotic breakdown

- Definition: Leakage of contents from surgical connection sites (external, drainage site, or imaging findings), including gastrointestinal, biliary, pancreatic ducts, etc., which may lead to fever, abscess, or organ failure

- Grading:

- Mild: Asymptomatic, only imaging findings

- Moderate: Requires treatment, no permanent damage

- Severe: Requires surgical intervention or causes organ dysfunction

4 Arrhythmia

- ECG-confirmed cardiac rhythm abnormalities

- Grading: Standard grading

5 Cardiac arrest

- Cessation of mechanical cardiac activity with no circulatory signs

- Diagnostic criteria: ECG or clinical examination

- No grading

6 Cardiogenic pulmonary oedema

- Alveolar fluid accumulation due to cardiac dysfunction

- Grading: Standard grading

7 Deep vein thrombosis (DVT)

- Newly formed thrombus in the venous system detected by ultrasound, venography, or CT imaging

- Grading: Standard grading

8 Delirium

- Each criterion met scores one point:

- Inattention

- Disorientation

- Hallucinations-delusions-psychosis

- Psychomotor agitation or retardation

- Inappropriate speech or mood

- Sleep/wake cycle disturbance or symptom fluctuation

- Delirium can be diagnosed when score reaches 4 points

- No grading

9 Gastrointestinal bleed

- Clear clinical or endoscopic evidence of gastrointestinal bleeding

- Grading: Standard grading

10 Infection, source uncertain

- Clinically suspected infection with undetermined source

- Meeting two or more of the following:

- Core temperature < 36°C or > 38°C

- White blood cell count > 12 × 10^9 L^-1 or < 4 × 10^9 L^-1

- Respiratory rate >20 breaths per minute or PaCO2 < 4.7 kPa (35 mmHg)

- Heart rate >90 beats per minute

- Grading: Standard grading

11 Laboratory confirmed bloodstream infection

- At least one of the following, unrelated to infection at other sites:

- Positive blood culture with exclusion of other infection sources

- Clinical symptoms (fever, chills, or hypotension) plus supportive indicators (at least one of the following):

- a. Common skin contaminants isolated from two or more blood cultures drawn at different times

- b. Common skin contaminants isolated from intravascular catheter blood culture with appropriate antimicrobial therapy initiated by physician

- c. Positive blood antigen test

- Grading: Standard grading

12 Myocardial infarction

- Elevated cardiac injury markers (such as TnT) with one of the following:

- Ischemic symptoms

- New ECG abnormalities (such as ST changes, left bundle branch block, or pathological Q waves)

- Regional wall motion abnormalities

- Grading: Standard grading

13 Myocardial injury after non-cardiac surgery (MINS)

- Definition: TnT ≥0.03 ng/mL without other causes (including renal dysfunction)

- No grading

14 Pneumonia

- Must meet all of the following conditions:

- Radiological criteria (requires two chest X-rays if underlying lung or heart disease, otherwise one is sufficient):

- New or progressive and persistent infiltrates

- Consolidation

- Cavitation

- Systemic manifestations (at least one):

- Unexplained fever >38°C

- White blood cell count <4×10^9/L or >12×10^9/L

- For patients ≥70 years old, unexplained mental status changes

- Respiratory-related changes (at least two):

- New purulent sputum or sputum character changes, or increased secretions/need for more suctioning

- New or worsening cough, dyspnea, tachypnea

- Rales or bronchial breath sounds

- Worsening gas exchange (hypoxemia, need for increased oxygen or ventilator support)

- Grading: Standard grading

15 Paralytic ileus

- Unable to tolerate diet or absence of flatus 3 days postoperatively

- Notes:

- Doctors may forget to record flatus or fasting status. If there are no relevant records at all, gastrointestinal symptoms (such as abdominal distension or vomiting) are required for diagnosis

- If fasting is due to enterocutaneous fistula, paralytic ileus should not be diagnosed unless there are gastrointestinal symptoms

- Grading: Standard grading

16 Postoperative haemorrhage

- Blood loss requiring transfusion or surgical hemostasis within 72 hours postoperatively

- Notes: If the patient has minimal postoperative drainage with light color but still receives transfusion, consider intraoperative bleeding or pre-existing anemia as the cause; do not diagnose postoperative hemorrhage

- Grading: Only includes moderate and severe; surgical hemostasis required is severe

17 Pulmonary embolism (PE)

- Newly formed thrombus in the pulmonary arterial system

- Grading: Standard grading

18 Stroke

- Persistent motor, sensory, or cognitive dysfunction due to embolic or hemorrhagic cerebrovascular events

- Grading: Standard grading

19 Surgical site infection (superficial)

- Infection within 30 days postoperatively

- Limited to incision skin and subcutaneous tissue

- Meeting any of the following:

- Purulent drainage from superficial incision

- Pathogenic microorganisms cultured from aseptically obtained fluid or tissue from superficial incision

- Incision site shows any of pain/tenderness, localized swelling, redness, or heat, and surgeon deliberately opens superficial incision (may be culture positive or not cultured, but culture negative does not meet criteria)

- Surgeon or attending physician diagnoses surgical site infection

- Grading: Standard grading

20 Surgical site infection (deep)

- Occurs within 30 days postoperatively if no implant, or within 1 year if implant present

- Infection involves deep soft tissues of incision (such as fascia and muscle layers)

- Patient meets at least one of the following:

- Purulent drainage from deep incision (not from organ/space)

- Deep incision spontaneously dehisces or is deliberately opened by surgeon, and patient has fever (>38°C) or local pain/tenderness, culture positive or not cultured (culture negative does not meet this criterion)

- Abscess or other evidence of infection found on direct examination, surgery, pathology, or imaging of deep incision

- Surgeon or attending physician diagnoses surgical site infection

- Grading: Standard grading

21 Surgical site infection (organ/space)

- Occurs within 30 days postoperatively

- Infection is surgery-related and involves body parts other than skin incision, fascia, muscle layers

- Patient meets at least one of the following:

- Purulent drainage from drain placed through small incision into organ/space

- Pathogenic microorganisms cultured from aseptically obtained organ/space fluid or tissue

- Abscess or other evidence of infection confirmed by direct examination, reoperation, pathology, or imaging of organ/space

- Surgeon or attending physician diagnoses organ/space surgical site infection

- Grading: Standard grading

22 Urinary tract infection

- Following two conditions appear 24h postoperatively:

- Positive urine culture (≥10^5 CFU/ml) with no more than two types of microorganisms

- Simultaneous occurrence of at least one of the following symptoms or signs: fever (>38°C), urgency, frequency, dysuria, suprapubic tenderness, costovertebral angle pain or tenderness, with no other recognized cause

- Grading: Standard grading

**### JSON Output Format**

1. First, output a think field containing an array, where each element is a string representing thoughts and considerations about the case. For example:

- "Patient recovered well postoperatively with no obvious discomfort"

- "Elevated troponin T suggests possible myocardial injury"

2. Then, output a complications field containing an array, where each element includes:

- name: Name of the complication, must be one of the 22 predefined complications

- grading: Severity grading, can only be one of the following four options:

- "Mild" - Mild: No treatment required

- "Moderate" - Moderate: Treatment required, no long-term effects

- "Severe" - Severe: Causes organ dysfunction or death, significantly prolongs hospital stay

- "Null" - No grading

Complete format example:

```json

{

"think": [

"Patient after kidney transplant surgery, acute kidney injury diagnosis excluded according to guidelines.",

"First postoperative troponin T elevated to 0.040 ng/mL, meeting definition of myocardial injury after non-cardiac surgery."

],

"complications": [

{

"name": "Myocardial Injury After Non-cardiac Surgery",

"grading": "Null"

}

]

}

```

Notes:

- If no postoperative complications are found, write "No postoperative complications" in the complications array

- If postoperative complications are found, they must be included in the above 22 complications; custom complications are prohibited

- Standard grading follows:

- Mild: No treatment required

- Moderate: Treatment required, no long-term effects

- Severe: Causes organ dysfunction or death, significantly prolongs hospital stay

**### Medical Record Data**

**{Same as the corresponding content in the previous section}**

#### Targeted version（Taking acute kidney injury as an example）

You are a senior surgeon. Your task is to determine whether the patient developed acute kidney injury postoperatively based on the medical record data. The diagnostic criteria for this complication are: **Acute Kidney Injury**

- Definition: Within seven days postoperatively, meeting KDIGO criteria:

- Grade I: Creatinine 1.5–1.9 times baseline or urine output reduction for 6–12 hours

- Grade II: Creatinine 2–2.9 times baseline or urine output reduction >12 hours

- Grade III: Creatinine ≥3 times baseline or requiring renal replacement therapy

- Notes:

- If the surgery is kidney transplantation, postoperative acute kidney injury should not be diagnosed

- If there are no preoperative creatinine test results, calculate based on baseline value of 84

Note: All diagnostic criteria must be met (unless the criteria explicitly state that meeting one of them is sufficient). Please output in the following JSON format:

```json

{

"think": "...(your step-by-step thought process)",

"bool": "True/False (determine whether the patient developed complications, can only output True or False)",

"grading": "Null/Mild/Moderate/Severe (severity of the complication, can only output one option)"

}

```

**### Medical Record Data**

**{Same as the corresponding content in the previous section}**

### Appendix methodology-Evaluation Metrics Calculation

#### Overview

This appendix provides detailed mathematical formulations for the evaluation metrics used in our perioperative complication detection study. All metrics are calculated using standard multi-class classification approaches with both micro-averaged and macro-averaged strategies.

#### Data Structure and Preprocessing

##### Label Representation

Each clinical case is represented as a set of complications detected. For a given case $i$: - $Y_{i}$ = Ground truth label set (gold standard) - $\hat{Y}_{i}$ = Predicted label set from model - $n$ = Total number of cases in the evaluation dataset

##### Label Extraction Process

From model outputs containing structured JSON or text responses, we extract complication labels using pattern matching and parsing algorithms. Each extracted label represents a specific perioperative complication according to the European Perioperative Clinical Outcome (EPCO) definitions.

#### Core Metrics Definition

##### True Positives, False Positives, and False Negatives

For each case $i$, we calculate: - **True Positives (TP)**: $TP_{i}=\left| Y_{i}\cap\hat{Y}_{i} \right|$ - **False Positives (FP)**: $FP_{i}=\left| \hat{Y}_{i}-Y_{i} \right|$ - **False Negatives (FN)**: $FN_{i}=\left| Y_{i}-\hat{Y}_{i} \right|$

Where: - $\left| S \right|$ denotes the cardinality (size) of set $S$ - $\cap$ denotes set intersection - $-$ denotes set difference - $Y_{i}\cap\hat{Y}_{i}$ represents correctly predicted complications for case $i$ - $\hat{Y}_{i}-Y_{i}$ represents incorrectly predicted complications (model predicted but not in ground truth) - $Y_{i}-\hat{Y}_{i}$ represents missed complications (in ground truth but not predicted by model)

#### Micro-Averaged Metrics

##### Calculation Method

Micro-averaging aggregates the contributions of all cases to compute global metrics:

1. **Global Aggregation**:

$$TP_{global}=\sum_{i=1}^{n} TP_{i}=\sum_{i=1}^{n} \left| Y_{i}\cap\hat{Y}_{i} \right|$$

$$FP_{global}=\sum_{i=1}^{n} FP_{i}=\sum_{i=1}^{n} \left| \hat{Y}_{i}-Y_{i} \right|$$

$$FN_{global}=\sum_{i=1}^{n} FN_{i}=\sum_{i=1}^{n} \left| Y_{i}-\hat{Y}_{i} \right|$$

2. **Micro-Averaged Precision**:

$$Precision_{micro}=\frac{TP_{global}}{TP_{global}+FP_{global}}$$

- Where:
  - $TP_{global}$ = Total correctly predicted complications across all cases
  - $FP_{global}$ = Total incorrectly predicted complications across all cases
  - $TP_{global}+FP_{global}$ = Total predicted complications across all cases
- **Micro-Averaged Recall**:

$$Recall_{micro}=\frac{TP_{global}}{TP_{global}+FN_{global}}$$

- Where:
  - $FN_{global}$ = Total missed complications across all cases
  - $TP_{global}+FN_{global}$ = Total actual complications across all cases
- **Micro-Averaged F1-Score**:

$$F1_{micro}=\frac{2\times Precision_{micro}\times Recall_{micro}}{Precision_{micro}+Recall_{micro}}$$

##### Edge Case Handling

- If $TP_{global}+FP_{global}=0$: $Precision_{micro}=0.0$
- If $TP_{global}+FN_{global}=0$: $Recall_{micro}=0.0$
- If $Precision_{micro}+Recall_{micro}=0$: $F1_{micro}=0.0$

#### Macro-Averaged Metrics

##### Calculation Method

Macro-averaging computes metrics for each case individually, then averages across all cases:

1. **Per-Case Metrics**: For each case $i$:

$$Precision_{i}=\frac{TP_{i}}{TP_{i}+FP_{i}}=\frac{\left| Y_{i}\cap\hat{Y}_{i} \right|}{\left| \hat{Y}_{i} \right|}$$

$$Recall_{i}=\frac{TP_{i}}{TP_{i}+FN_{i}}=\frac{\left| Y_{i}\cap\hat{Y}_{i} \right|}{\left| Y_{i} \right|}$$

$$F1_{i}=\frac{2\times Precision_{i}\times Recall_{i}}{Precision_{i}+Recall_{i}}$$

2. **Macro-Averaged Metrics**:

$$Precision_{macro}=\frac{1}{n}\sum_{i=1}^{n} Precision_{i}$$

$$Recall_{macro}=\frac{1}{n}\sum_{i=1}^{n} Recall_{i}$$

$$F1_{macro}=\frac{1}{n}\sum_{i=1}^{n} F1_{i}$$

##### Edge Case Handling for Individual Cases

- If $TP_{i}+FP_{i}=0$ (no predictions for case $i$): $Precision_{i}=0.0$
- If $TP_{i}+FN_{i}=0$ (no ground truth labels for case $i$): $Recall_{i}=0.0$
- If $Precision_{i}+Recall_{i}=0$: $F1_{i}=0.0$

#### Variable Definitions Summary

| Variable | Definition |
| --- | --- |
| $Y_{i}$ | Ground truth label set for case $i$ |
| $\hat{Y}_{i}$ | Predicted label set for case $i$ |
| $n$ | Total number of cases |
| $TP_{i}$ | True positives for case $i$ |
| $FP_{i}$ | False positives for case $i$ |
| $FN_{i}$ | False negatives for case $i$ |
| $TP_{global}$ | Sum of true positives across all cases |
| $FP_{global}$ | Sum of false positives across all cases |
| $FN_{global}$ | Sum of false negatives across all cases |
| $Precision_{i}$ | Precision for individual case $i$ |
| $Recall_{i}$ | Recall for individual case $i$ |
| $F1_{i}$ | F1-score for individual case $i$ |
| $Precision_{micro}$ | Micro-averaged precision |
| $Recall_{micro}$ | Micro-averaged recall |
| $F1_{micro}$ | Micro-averaged F1-score |
| $Precision_{macro}$ | Macro-averaged precision |
| $Recall_{macro}$ | Macro-averaged recall |
| $F1_{macro}$ | Macro-averaged F1-score |

#### Implementation Details

##### Code Implementation

The evaluation metrics are implemented in Python using the following functions:

def evaluate_micro(golds, preds):
 """
 Calculate micro-averaged Precision, Recall, F1.

 Args:
 golds: List of ground truth label sets
 preds: List of predicted label sets

 Returns:
 precision, recall, f1 (float values)
 """
 TP_global = FP_global = FN_global = 0

 for gold, pred in zip(golds, preds):
 gold_set = set(gold)
 pred_set = set(pred)
 TP_global += len(gold_set & pred_set)
 FP_global += len(pred_set - gold_set)
 FN_global += len(gold_set - pred_set)

 precision = TP_global / (TP_global + FP_global) if (TP_global + FP_global) else 0.0
 recall = TP_global / (TP_global + FN_global) if (TP_global + FN_global) else 0.0
 f1 = 2 * precision * recall / (precision + recall) if (precision + recall) else 0.0

 return precision, recall, f1

def evaluate_macro_sample(golds, preds):
 """
 Calculate macro-averaged Precision, Recall, F1.

 Args:
 golds: List of ground truth label sets
 preds: List of predicted label sets

 Returns:
 precision, recall, f1 (float values)
 """
 p_list, r_list, f_list = [], [], []

 for gold, pred in zip(golds, preds):
 gold_set = set(gold)
 pred_set = set(pred)
 tp = len(gold_set & pred_set)
 fp = len(pred_set - gold_set)
 fn = len(gold_set - pred_set)

 precision = tp / (tp + fp) if (tp + fp) else 0.0
 recall = tp / (tp + fn) if (tp + fn) else 0.0
 f1 = 2*precision*recall/(precision+recall) if (precision+recall) else 0.0

 p_list.append(precision)
 r_list.append(recall)
 f_list.append(f1)

 macro_p = sum(p_list)/len(p_list) if p_list else 0.0
 macro_r = sum(r_list)/len(r_list) if r_list else 0.0
 macro_f1 = sum(f_list)/len(f_list) if f_list else 0.0

 return macro_p, macro_r, macro_f1

#### Metric Interpretation

##### Micro vs. Macro Averaging

- **Micro-averaging** gives equal weight to each complication detection across the entire dataset, making it more sensitive to performance on frequently occurring complications
- **Macro-averaging** gives equal weight to each case, making it more sensitive to performance consistency across cases regardless of the number of complications per case

##### Clinical Relevance

- **Precision** measures the proportion of predicted complications that are actually present (low false positive rate)
- **Recall** measures the proportion of actual complications that are correctly detected (low false negative rate)
- **F1-score** provides a balanced measure combining both precision and recall, particularly important in clinical contexts where both missing complications (false negatives) and false alarms (false positives) have significant consequences

#### Primary Metric Selection

F1-score serves as our primary performance indicator due to its balanced consideration of both precision and recall, which is particularly important in clinical applications where both types of errors can impact patient care and clinical decision-making.
